# Tracking ℛ of COVID-19: A New Real-Time Estimation Using the Kalman Filter

**DOI:** 10.1101/2020.04.19.20071886

**Authors:** Francisco Arroyo-Marioli, Francisco Bullano, Simas Kučinskas, Carlos Rondón-Moreno

**Author notes:** We would like to thank Christiane Baumeister, Ralph Brinks, Eric Budish, Ricardo Hausmann, Artūras Juodis, Jan Keil, Siem Jan Koopman, Andrés Neumeyer, Mathieu Pedemonte, Sergio Ocampo-Díaz, Sándor Sóvágó, Eduardo Undurraga, Iván Werning, as well as seminar participants at the Central Bank of Chile and Harvard Growth Lab for their comments and suggestions. We also thank Chen Lin for excellent research assistance. Replication files are available here: online. The views and conclusions presented in this paper are exclusively those of the authors and do not necessarily reflect the position of the Central Bank of Chile or of the Board members. The present paper subsumes the previous working papers of Arroyo-Marioli, Bullano and Rondón-Moreno (2020) and Kučinskas (2020).

## Abstract

We develop a new method for estimating the effective reproduction number of an infectious disease (ℛ) and apply it to track the dynamics of COVID-19. The method is based on the fact that in the SIR model, ℛ is linearly related to the growth rate of the number of infected individuals. This time-varying growth rate is estimated using the Kalman filter from data on new cases. The method is easy to implement in standard statistical software, and it performs well even when the number of infected individuals is imperfectly measured, or the infection does not follow the SIR model. Our estimates of ℛ for COVID-19 for 124 countries across the world are provided in an interactive [online dashboard](https://bit.ly/Rtlive), and they are used to assess the effectiveness of non-pharmaceutical interventions in a sample of 14 European countries.

## 1 Introduction

The effective reproduction number (ℛ) plays a central role in the epidemiology of infectious diseases. ℛ is defined as the average number of secondary cases produced by a primary case (Wallinga and Teunis, 2004; Nishiura and Chowell, 2009; Cori et al., 2013). The effective reproduction number varies over time, due to the depletion of susceptible individuals as well as changes in other factors, including control measures, contact rates, and climatic conditions. The *basic reproduction number*, denoted by ℛ_0_, measures the average number of secondary cases produced by a primary case when the population is fully susceptible (Dietz, 1993; Chowell and Brauer, 2009). Analogously to the effective reproduction number, the basic reproduction number is also affected by multiple variables (Delamater et al., 2019).

In standard models, the number of infected individuals increases as long as ℛ > 1. Real-time estimates of ℛ are therefore essential for public policy decisions during a pandemic (Atkeson, 2020; Leung, 2020). Such estimates can be used to study the effectiveness of non-pharmaceutical interventions (NPIs), or assess what fraction of the population needs to be vaccinated to reach herd immunity (Chinazzi et al., 2020; Kucharski et al., 2020; Wang et al., 2020). Some social scientists have argued that ℛ < 1 should be viewed as a fundamental constraint on public policy during the current COVID-19 pandemic (Budish, 2020).

In this paper we develop a new method to estimate ℛ in real time. The method exploits the fact that in the benchmark SIR model, ℛ is linearly related to the growth rate of the number of infected individuals (Kermack and McKendrick, 1927). Our estimation procedure consists of three steps. First, we use data on new cases to construct a time series of how many individuals are infected at a given point in time. Then, we estimate the growth rate of this time series with the Kalman filter. In the final step, we leverage the theoretical relationship given by the SIR model to obtain ℛ from the estimated growth rate. We show theoretically that the estimates are not sensitive to potential model misspecification, and they are fairly accurate even when new cases are imperfectly measured.

We apply our methodology to estimate the ℛ of COVID-19 in real-time. Our estimates for 124 countries across the world are provided in an online dashboard and can be explored interactively (link to dashboard). In empirical applications, we use these estimates to calculate the basic reproduction number (ℛ_0_) and evaluate the effects of NPIs in reducing ℛ for a sample of 14 European countries.

Under our baseline assumption that the serial interval for COVID-19 is seven days, we estimate the basic reproduction number (ℛ_0_) to be 2.66 (95% CI: 1.98–3.38). Next, we find that lockdowns, measures of self-isolation, and social distancing all have a statistically significant effect on reducing ℛ. However, we also demonstrate the importance of accounting for voluntary changes in behavior. In particular, we document that most of the decline in mobility in our sample happened before the introduction of lockdowns. Failing to account for voluntary changes in behavior leads to substantially over-estimated effects of NPIs.

### Related Literature

There are two broad classes of methods that can be used to estimate ℛ in real time (Chowell and Brauer, 2009; Nishiura and Chowell, 2009; Gostic et al., 2020). First, one can estimate a fully-specified epidemiological model and then construct a model-implied time series for ℛ (Chowell et al., 2007; Cazelles et al., 2018; Dehning et al., 2020; Kucharski et al., 2020). Second, one may use approaches that leverage information on the serial interval of a disease (i.e., time between onset of symptoms in a case and onset of symptoms in his/her secondary cases (Wallinga and Teunis, 2004; Wallinga and Lipsitch, 2007; Cori et al., 2013)). For example, imagine a disease with a fixed serial interval of, say, three days. In that case, we could estimate ℛ by simply dividing the number of new cases today by the number of new cases three days ago. Cori et al. (2013) exploit this idea to develop a Bayesian estimator that accounts for the randomness in the onset of infections as well as variation in the serial interval; see also Thompson et al. (2019). This method is implemented in a popular ℛ package `EpiEstim`.

The method proposed in this paper attempts to strike a balance between the two approaches mentioned above. Although our estimator is derived from standard epidemiological theory, we use the smallest amount of theoretical structure that is necessary to obtain our estimator. In particular, the theoretical relationship used to derive our estimator is exactly valid not only in the standard SIR model with constant parameters, but also in the SIS model and a generalized SIR model with time-varying parameters and stochastic shocks. Relative to the existing literature, our estimator does not need any statistical tuning parameters, and it does not require parametric assumptions on the distribution of new cases (such as assuming that new cases are Poisson distributed). For example, the method of Cori et al. (2013) assumes that ℛ is constant over fixed windows of duration *τ*; *τ* effectively becomes a tuning parameter that needs to be chosen by the user. Our approach and its mathematical derivation share some similarities with the estimator proposed by Bettencourt and Ribeiro (2008).

A key advantage of using the Kalman filter for estimating ℛ is that valid confidence bounds are readily obtained. Explicitly accounting for the dynamics in ℛ via the state equation ensures that the estimated effective reproduction numbers are not excessively volatile, with the optimal amount of filtering estimated from the data. In addition, the Kalman smoother allows the researcher to use full-sample information efficiently when estimating ℛ. Finally, our method can be used with both classical and Bayesian techniques, as we demonstrate in the empirical application.

## 2 A Real-Time Estimator

We now derive our estimator for the SIR model (Kermack and McKendrick, 1927). In the Appendix, we show that we can obtain the same estimator from an SIS model (Appendix A.1), and an SIR model with stochastic shocks (Appendix A.2).

### 2.1 Deriving the Estimator

The standard SIR model in discrete time describes the evolution of susceptible (*S*_*t*_), infected (*I*_*t*_), and recovered (ℛ_*t*_) individuals by the following equations (Allen and Van Den Driessche, 2008; Stock, 2020):

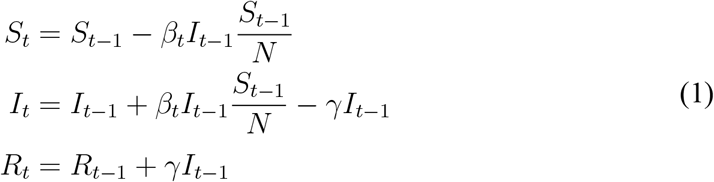

The model is stated at a daily frequency. Here, *N* ≡ *S*_*t*_ + *I*_*t*_ + *R*_*t*_ is the population size, *β*_*t*_ is the daily transmission rate, and *γ* is the daily transition rate from infected to recovered. The recovered group consists of individuals who have either died or fully recovered. We allow the transmission rate *β*_*t*_ to vary over time. For example, individuals may choose to to reduce their social interactions voluntarily, or they could be subject to government policy restrictions.

The *basic reproduction number*, 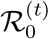, is defined as 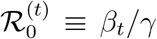, and it gives the average number of individuals infected by a single infected individual when everyone else is susceptible. Since the transmission rate *β*_*t*_ varies over time, the basic reproducetion number is generally time varying as well. The *effective reproduction number*, ℛ_*t*_, is defined as 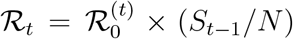, and it equals the average number of individuals infected by a single infected individual when a fraction (*S*_*t*−1_/*N*) of individuals is susceptible.

From Eq. (1) the daily growth rate in the number of infected individuals is

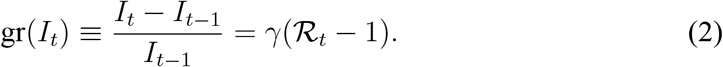

Denoting the estimated growth rate of infected individuals by 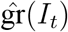, and given a value for the transition rate *γ*, the plug-in estimator for the effective reproduction number is

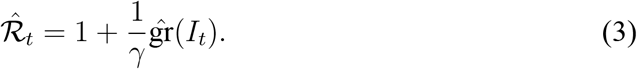

For the estimator to be feasible, we need to (i) calibrate the transition rate from infectious to recovered, *γ*; and (ii) estimate the growth rate of *I*_*t*_. There are two potential strategies for choosing *γ*. First, we can use external medical evidence given that *γ*^−1^ is the average infectious period. Second, information on the serial interval of the disease can be employed, given that the serial interval in the SIR model also equals *γ*^−1^ (Ma, 2020).

To estimate the growth rate of *I*_*t*_ empirically, we first construct a time series for *I*_*t*_ from data on new cases. The SIR model in Eq. (1) implies that

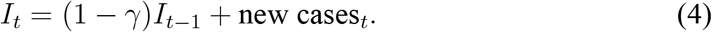

We initialize *I*_*t*_ by *I*_0_ = *C*_0_ where *C*_0_ is the total number of infectious cases at some initial date, and then construct subsequent values of *I*_*t*_ recursively.

Given the time series for *I*_*t*_, we use standard Kalman-filtering tools to smooth the observed growth rate of *I*_*t*_. In particular, we specify the following state-space model for the growth rate of *I*_*t*_:

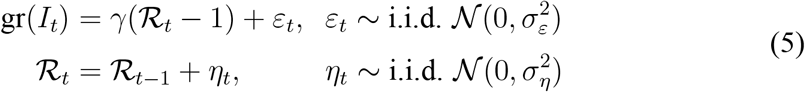

We estimate ℛ_*t*_ by the Kalman smoother (see Durbin and Koopman, 2012, Chapter 2). The Kalman smoother provides optimal estimates of ℛ_*t*_ (in the sense of minimizing mean-squared error) given the full-sample information on gr(*I*_*t*_), provided that the data are generated by the model in Eq. (5).

To estimate the unknown parameters 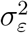 and 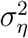 in Eq. (5), both classical and Bayesian methods can be used. However, sample sizes are usually limited in practice, especially early on in the epidemic. Hence, incorporating prior knowledge generally leads to better-behaved estimates. The state-space model above—also known as the local-level model—can also be thought as a model-based version of exponentially-weighted moving-average smoothing (Muth, 1960).

The state-space model in Eq. (5) can be viewed as a reduced-form time-series specification. The local-level model can capture fairly rich dynamic patterns in the data (Commandeur and Koopman, 2007; Durbin and Koopman, 2012). In addition, in Appendix A.3, we provide a theoretical rationale for the local-level specification. In particular, Eq. (5) arises naturally in the SIR model (in the early stages of an epidemic) when the transmission rate *β*_*t*_ follows a random walk.

From Eq. (4), the growth rate gr(*I*_*t*_) is bounded below by (−*γ*). Hence, for any estimator of gr(*I*_*t*_) that is some weighted average of the observed growth rates, the point estimate of ℛ_*t*_ is automatically non-negative. To ensure that lower confidence bounds are positive as well, we estimate the *q*-th quantile of ℛ_*t*_ by 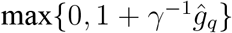, where 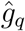 is an estimate of the *q*-th quantile of gr(*I*_*t*_). In addition (see Appendix A.4), our empirical estimates remain similar when we use a modified version of the Carter and Kohn (1994) algorithm which discards random draws violating the non-negativity constraint. Alternatively, it is possible to avoid this type of truncation by using non-linear filtering methods (Creal, 2012).

### 2.2 Sensitivity to Model Misspecification and Data Problems

Tracking the evolution of ℛ_*t*_ is notoriously difficult. Human-contact dynamics, testing, and changes in case definitions affect the flow and quality of the available information. In this section, we test the sensitivity of our estimator to two notable issues: (i) model misspecification; and (ii) data problems (such as reporting delays or imperfect detection of infectious individuals).

For the first issue, model misspecification, a natural concern is whether the true dynamics of the disease are well captured by the benchmark SIR model. We address this issue in two ways. First, we show that our estimator remains exactly valid in the SIS model in which individuals do not obtain immunity (Appendix A.1) and a generalized SIR model with stochastic shocks (Appendix A.2). In addition, provided that the average duration of infectiousness is correctly specified, we find that our estimator yields accurate results even when the true model is SEIR rather than SIR (Appendix A.5). Second, we note that the error term *ε*_*t*_ in the state-space model described by Eq. (5) can be interpreted as model error. Therefore, our estimates as well as their confidence intervals explicitly account for (some amount of) potential misspecification.

The second issue relates to data reliability. For COVID-19, testing constraints and high asymptomatic prevalence (Arons et al., 2020; Nishiura et al., 2020a; Streeck et al., 2020), in particular, make it challenging to identify all infectious individuals. The simplicity of our estimator allows us to analytically characterize the effects of potential measurement error (see Appendix A.6). Furthermore, we use these results to investigate the quantitative performance of the estimator in a number of empirically relevant underdetection scenarios using Monte Carlo simulations. Overall, we conclude that our method provides accurate estimates in all cases that we analyze.

## 3 Estimates for COVID-19

We now use data from the John Hopkins CSSE repository to obtain real-time estimates of ℛ_*t*_ for COVID-19 (Dong et al., 2020). In our estimations, we include all countries for which we have at least 20 daily observations after the cumulative number of confirmed COVID-19 cases reaches 100. Our sample period starts on 2020-01-23 and finishes on 2020-05-06. Appendix A.7 describes the details of the estimation procedure. Table A.5 in the Appendix contains the GATHER checklist (Stevens et al., 2016), summarizing the details of the analysis.

For the baseline estimates, we assume that people are infectious for *γ*^−1^ = 7 days on average, similarly to Maier and Brockmann (2020) and Prem et al. (2020). This assumption is consistent with the evidence on the serial interval of COVID-19. For example, Flaxman et al. (2020) use an average serial interval of 6.5 days. Recent studies find that estimates of the serial interval for COVID-19 generally range between 4 and 9 days (Nishiura et al., 2020b; Park et al., 2020; Sanche et al., 2020). In addition, we document that *γ*^−1^ = 7 leads to estimates of the basic reproduction number (ℛ_0_) that are in line with the recent estimates in the literature (Liu et al., 2020). However, we also investigate the effects of different choices for *γ* on our results. In general, by virtue of Eq. (3), changing *γ* tilts the estimates of ℛ_*t*_ around one, with higher values of the serial interval pushing the estimates away from one and lower values pushing the estimates towards one. For example, if 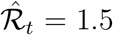 for *γ*^−1^ = 7 days, increasing the serial interval to 8 days increases the estimate to 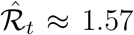. Conversely, if 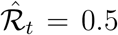 for *γ*^−1^ = 7 days, increasing the serial interval to 8 days decreases the estimate to 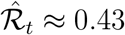.

In Appendix A.8, we perform two empirical validation exercises of our estimates. First, we document that our estimates of ℛ_*t*_ are predictive of future deaths. Given that deaths are arguably more accurately measured, this finding alleviates concerns regarding potential data reliability issues that could contaminate our estimates. Second, we find that past mobility data is predictive of future values of ℛ_*t*_. In Appendix A.10, we additionally compare our estimates to those obtained using the method of Cori et al. (2013). We find that our estimates are highly correlated to the estimates produced by the Cori et al method, with the average correlation coefficient across different countries equal to 0.80 (median: 0.89). Jointly, these exercises suggest that our estimates contain valuable information on the dynamics of COVID-19.

### 3.1 Estimated Effective Reproduction Numbers

Our estimates of ℛ_*t*_ for selected countries are shown in Figure 1. Estimates for the remaining countries can be found in the associated online dashboard.

**Figure 1.**
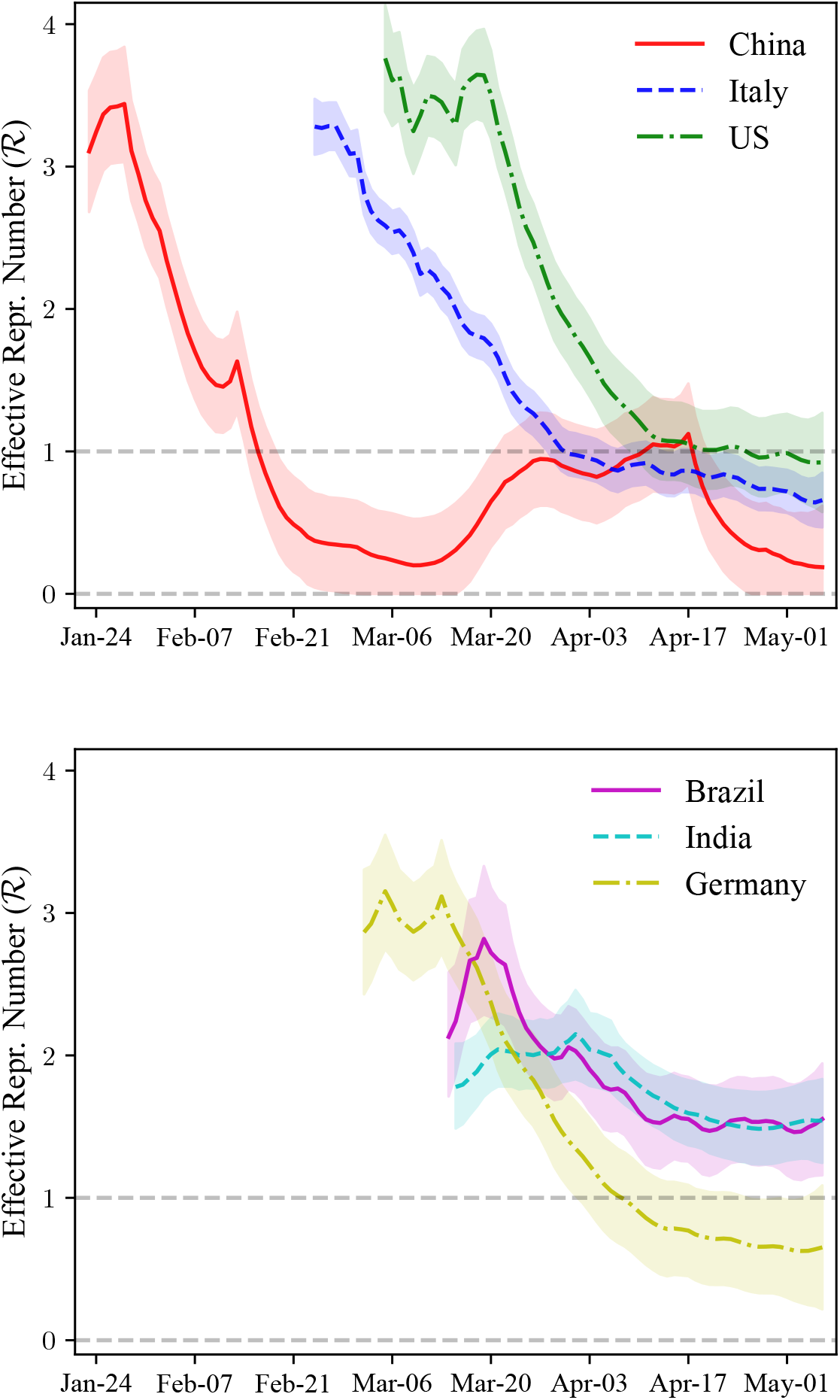
ℛ_*t*_ of COVID-19: Selected Countries. *Notes*: Estimates of the effective reproduction number (ℛ_*t*_) of COVID-19 for selected countries. For each country, the sample consists of all dates after the total number of reported cases has reached 100. 65% credible bounds shown by the shaded areas. *Top panel*: estimated effective reproduction numbers for China, Italy, and the US. *Bottom panel:* estimated effective reproduction numbers for Brazil, India, and Germany.

The top panel of Figure 1 plots the estimated effective reproduction numbers for China, Italy, and the US. In the Appendix (Figure A.7), we also provide a graph of the raw data on the growth rate of the number of infected individuals that is used for estimating ℛ_*t*_. For all three countries, the estimated ℛ_*t*_ is initially above 3. For China, the estimated ℛ_*t*_ declined rapidly, falling below one around the third week of February. According to our estimates, ℛ_*t*_ in China fell below one 24 days after the beginning of the epidemic in the country (with the start of the epidemic defined as reaching 100 cumulative confirmed cases of COVID-19). However, the estimated ℛ_*t*_ in China drifted up towards one during late March and early April, potentially caused by a wave of imported cases. Note that there is an upwards jump in the estimated ℛ_*t*_ for China around the second week of February. This jump was caused by a temporary change in COVID-19 case definitions in the Hubei province in China; the new definition included clinically-diagnosed COVID-19 cases (Tsang et al., 2020).

In Italy, the estimated ℛ_*t*_ fell steadily since March but at a slower rate than previously observed in China, with the point estimate for Italy falling below one in early April. Our estimates indicate that it took 36 days for ℛ_*t*_ to fall below one after the start of the epidemic in Italy. In the US, the point estimates of ℛ_*t*_ were fairly flat in the first two weeks of the epidemic, hovering around 3.5. We note, however, that it is likely that the fraction of non-detected cases in the US went down substantially in this period, inflating the estimates of ℛ_*t*_ upward (see Appendix A.6). In particular, the daily number of tests conducted in the U.S went up dramatically during this period, increasing by a factor of 45 between March 8, 2020 and March 25, 2020 (Our World in Data, 2020). It took 52 days for the estimated ℛ_*t*_ to fall below one for the first time in the US after the start of the epidemic, or more than twice as long as in China. The point estimate of ℛ_*t*_ in the US at the end of the sample is below one and equal to 0.92 (95% CI: 0.17–1.66).

The bottom panel of Figure 1 plots the estimated effective reproduction numbers for Brazil, India, and Germany. The pattern observed in Germany is similar to that previously seen in Italy and the United States. The estimated ℛ_*t*_ in Germany falls below one 37 days after the beginning of the pandemic, almost identically to Italy. In Brazil and India, the point estimates of ℛ_*t*_ are lower at the beginning of the pandemic than in the other countries plotted here. The effective reproduction numbers at the beginning of the epidemic are estimated to be 2.13 (95% CI: 0.81–3.04) in Brazil, and 1.78 (95% CI: 0.92–2.41) in India. In contrast, for example, ℛ_*t*_ is estimated to be 2.86 (95% CI: 1.91– 3.81) in Germany at the beginning of the pandemic. We emphasize that the estimated confidence bounds are wide, indicating substantial uncertainty about the true values of ℛ_*t*_. Hence, substantial caution must be exercised when comparing the estimates of ℛ_*t*_ across countries and over time.

A natural concern with any estimator of ℛ_*t*_ applied to COVID-19 is that the estimator may be biased if only a fraction of all COVID-19 cases is detected. In the Appendix (Appendix A.6), we study the performance of our estimator under various assumptions on the reporting of COVID-19 cases. We show analytically that our estimator remains exactly valid even when only a fraction of all cases is detected (e.g., 10% of all cases are detected), provided that the fraction of all cases detected is constant over time. The estimates are also accurate under some other cases of misreporting. However, if the fraction of detected COVID-19 cases changes a lot over short windows of time, the estimator is biased. Finally, we investigate the performance of our estimator in a number of additional cases of imperfect reporting (such as a ramp-up in testing) that may be important in practice using Monte Carlo simulations. Overall, we conclude that our estimator is robust to potential mismeasurement of COVID-19 cases in a number of empirically-relevant scenarios.

In the Appendix (Figure A.8), we illustrate the difference between estimates of ℛ_*t*_ for China obtained by the Kalman smoother—as in our baseline estimation—and the Kalman filter. Intuitively, the Kalman smoother uses information from the full sample when estimating ℛ_*t*_, while the Kalman filter only uses information up to and including time *t* (Durbin and Koopman, 2012). As seen in the graph, while the two sets of estimates are fairly similar, the filtered estimates are substantially more volatile. In addition, the filtered estimates generally have wider credible bounds. As should be the case, the filtered and smoothed estimates are identical at the endpoint of the sample. From the perspective of epidemiological theory, the Kalman filter essentially produces what Fraser (2007) refers to as the instantaneous reproduction number, while the Kalman smoother yields the case reproduction number. The estimator proposed in the present paper therefore allows researchers to estimate the two types of reproduction numbers in a single unified framework.

In Figure A.8 in the Appendix, we also demonstrate the difference between our Bayesian estimates of ℛ_*t*_ and classical estimates obtained via maximum likelihood. For China, the two sets of estimates are virtually indistinguishable, indicating that the chosen priors have a small effect on the estimates. Of course, for other some countries in our sample, the data are less informative, and hence the priors have a more pronounced effect.

### 3.2 Basic Reproduction Number

We now use our estimates of ℛ_*t*_ to measure the basic reproduction number (ℛ_0_), i.e., the average number of individuals infected by a single infectious individual when the population is fully susceptible. We estimate ℛ_0_ by the average value of ℛ_*t*_ in the first week of the epidemic.

Table 1 shows the results for a sample of 14 European countries (Austria, Belgium, Denmark, France, Germany, Greece, Italy, Netherlands, Norway, Portugal, Spain, Sweden, Switzerland, and United Kingdom), as in Flaxman et al. (2020). Under our baseline assumption that the individuals are infectious for 7 days on average (*γ* = 1/7), we obtain an estimate of ℛ_0_ = 2.66 (95% CI: 1.98–3.38). For COVID-19, a recent meta-study has estimated a median ℛ_0_ of 2.79 (Liu et al., 2020), suggesting that our results are consistent with the current consensus estimates.

**Table 1.** Estimates of the Basic Reproduction Number (ℛ_0_)

| Number of Days Infectious: | 5 | 6 | 7 | 8 | 9 | 10 |
| --- | --- | --- | --- | --- | --- | --- |
| $\hat{\mathcal{R}}_0$ | 2.07 | 2.35 | 2.66 | 2.89 | 3.10 | 3.29 |
| CI Lower Bound (95%) | 1.51 | 1.75 | 1.98 | 2.17 | 2.37 | 2.54 |
| CI Upper Bound (95%) | 2.67 | 3.01 | 3.38 | 3.67 | 3.86 | 4.06 |

*Notes:* Estimates of the basic reproduction number ( $\mathcal{R}_0$ ) for a sample of 14 European countries. The countries included in the sample are Austria, Belgium, Denmark, France, Germany, Greece, Italy, Netherlands, Norway, Portugal, Spain, Sweden, Switzerland, and United Kingdom. The basic reproduction number is calculated by averaging our estimates of the effective reproduction number in the first 7 days of the epidemic, where the start of the epidemic is defined as the day when the cumulative number of cases reaches 100.

Table 1 also provides the estimated ℛ_0_ under different assumptions on the duration of infectiousness (or, equivalently in the SIR model, the average serial interval). As expected, the median estimate is sensitive to the choice of *γ*; we find an additional day of infectiousness increases ℛ_0_ by around 0.3.

### 3.3 Assessing Non-Pharmaceutical Interventions

Finally, we use our estimates to assess the effects of non-pharmaceutical interventions (NPIs) in the same sample of 14 European countries as in Section 3.2. We study a total of five NPIs: (i) lockdowns; (ii) bans of public events; (iii) school closures; (iv) mandated self-isolation when exhibiting symptoms; and (e) social distancing measures. We adopt the definitions of NPIs and their introduction dates provided by Flaxman et al. (2020).

We first perform an event-study exercise, inspired by event studies commonly used in economics and finance (MacKinlay, 1997). In this exercise, we compare the dynamics of the effective reproduction number before and after the introduction of a particular control measure. If the control measure is effective, we expect to observe a difference in the behavior of ℛ_*t*_ after its introduction. The difference may appear as either a change in levels (“jump”) or a change in trends (“kink”); the latter possibility is more likely in the present empirical context. This simple before-versus-after comparison is not free of potential bias. In particular, the comparison implicitly assumes that the behavior of ℛ_*t*_ before the intervention provides a good counterfactual for the (unobserved) future behavior of ℛ_*t*_ in the absence of the intervention. Nevertheless, we find this exercise instructive as a preliminary step in our analysis.

Figure 2a plots the estimated values of ℛ_*t*_ one week before and three weeks after the introduction of a lockdown. Since Sweden did not have a lockdown in the sample period considered, the figure is constructed using data from 13 countries. ℛ_*t*_ declines substantially after a lockdown is introduced, going from 2.11 (95% CI: 1.84–2.38) on the day of the intervention to 0.99 (95% CI: 0.87–1.11) three weeks later. However, ℛ_*t*_ is decreasing before the lockdown as well. In particular, there is no visually detectible break in the slope of ℛ_*t*_ in the three-week period after the introduction of the lockdown (i.e., no “kink”). In the Appendix, we show that other NPIs follow a similar pattern. In particular, we document the behavior of ℛ_*t*_ around the introduction of public-event bans (Figure A.10), case-based measures (such as self-isolation whenever feeling ill and experiencing fever; Figure A.11), school closures (Figure A.12), and social-distancing measures (Figure A.13). Except for school closures and public-event bans, there is no visually apparent break in the trend of ℛ_*t*_ around the date of the policy intervention.

**Figure 2.**
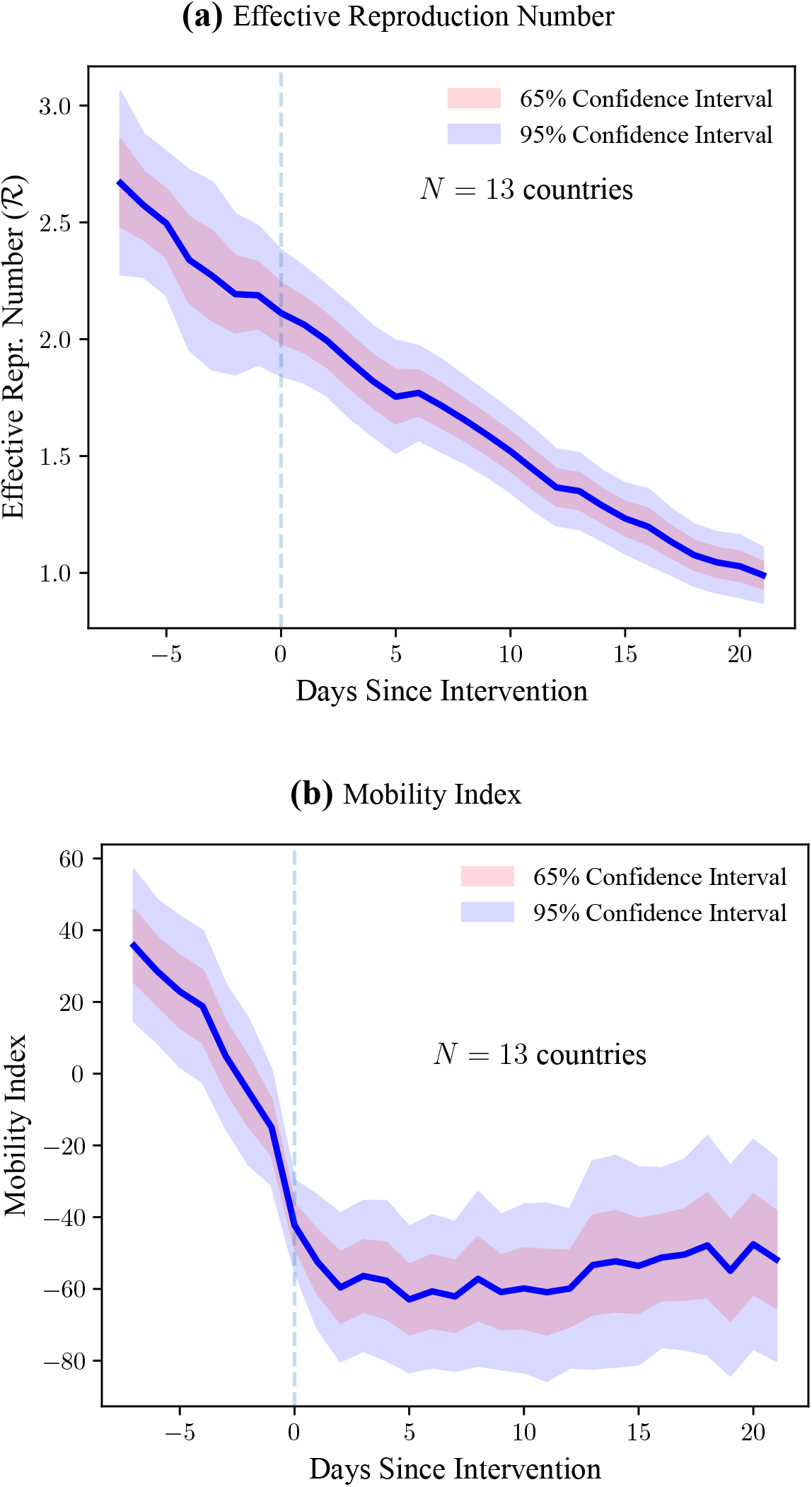
Event Study: ℛ and Mobility Around Lockdowns. *Notes*: *Top panel*: estimated effective reproduction number (ℛ_*t*_) one week before and three weeks after a lockdown is introduced in a country. *Bottom panel:* mobility index (constructed from “COVID-19 Community Mobility Reports” of Google (2020)) one week before and three weeks after a lockdown is introduced in a country. See Appendix A.8 for details on the construction of the mobility index. The original sample consists of 14 European countries studied by Flaxman et al. (2020). Heteroskedasticity-robust confidence bounds are shown by the shaded areas.

To further investigate the behavior of ℛ_*t*_ in the four-week window around lockdowns, we use mobility data from Google’s “COVID-19 Community Mobility Reports” (Google, 2020). Google uses smartphone location data to measure changes in mobility (relative to pre-pandemic levels) for six six types of places: (i) groceries and pharmacies; (ii) parks; (iii) transit stations; (iv) retail and recreation; (v) residential; and (vi) workplaces. Since these measures are strongly correlated, we take the first principal component of the six time series to construct an overall mobility index. The first principal component explains 83.03% of the total variation in Google’s mobility data.

Figure 2b shows that most of the decline in mobility occurs *before* the imposition of the lockdown, and remains low *thereafter*. This finding shows a clear change in people’s behavior in the early days of the pandemic. Shifting habits before the introduction of NPIs is consistent with the existence of private motives that can induce a reduction in mobility as people avoid becoming infected (Farboodi et al., 2020; Guerrieri et al., 2020; Krüger et al., 2020). Our results are also consistent with empirical evidence for the U.S and anecdotal reports from Sweden (Farboodi et al., 2020; The New York Times, 2020). The documented relationship between ℛ_*t*_ and mobility does not necessarily constitute evidence against the effectiveness of lockdowns. On the contrary, it is possible that lockdowns reinforce attitudes towards disease-awareness and self-isolation, helping to ensure lower values of ℛ_*t*_ in the long run.

A potential concern with the evidence in Figure 2a is that our estimates of ℛ_*t*_ use information from the full sample. Hence, estimates of ℛ_*t*_ *before* the lockdown implicitly depend on the estimates of ℛ_*t*_ *after* the lockdown. This feature of the estimation procedure may result in low statistical power to detect any effects of NPIs. To investigate this possibility, we conduct a power analysis (Appendix A.9). Given our empirical estimates of signal-to-noise ratios, we find that the statistical procedure appears sufficiently powerful to detect moderate changes in ℛ_*t*_.

To assess the effects of NPIs more formally, we employ the following fixed-effect regressions (Table 2). Specifically, we regress ℛ_*t*_ on a set of indicator variables capturing interventions and different types of fixed effects:

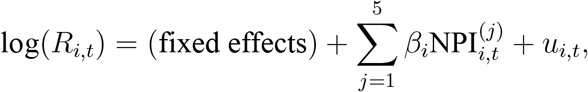

where *u*_*i,t*_ denotes the stochastic error term of the regression. The NPI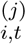 is an indicator variable that equals 1 after the *j*-th NPI is introduced, and zero before its introduction. The index *i* denotes countries, and *t* stands for the number of days since the outbreak of the epidemic.

**Table 2.** Effective Reproduction Number After Introduction of NPIs.

|  | *Dependent Variable: $\log(R_t)$* |  |  |  |
| --- | --- | --- | --- | --- |
|  | (1) | (2) | (3) | (4) |
| Lockdown | -0.65***<br>(0.04) | -0.13**<br>(0.05) | -0.1**<br>(0.05) | -0.04<br>(0.06) |
| Public Events | -0.09*<br>(0.04) | 0.22***<br>(0.04) | 0.21***<br>(0.04) | 0.36***<br>(0.08) |
| School Closure | -0.02<br>(0.06) | -0.19***<br>(0.05) | -0.18***<br>(0.06) | -0.1<br>(0.08) |
| Self Isolation | -0.13**<br>(0.06) | -0.09**<br>(0.05) | -0.04<br>(0.05) | 0.01<br>(0.08) |
| Social Distancing | -0.15**<br>(0.06) | -0.08<br>(0.06) | -0.11*<br>(0.06) | -0.14*<br>(0.08) |
| $N$ | 855 | 855 | 848 | 488 |
| $R^2$ | 0.53 | 0.87 | 0.88 | 0.91 |
| Country FE | ✓ | ✓ | ✓ | ✓ |
| Days-Since-Outbreak FE |  | ✓ | ✓ | ✓ |
| Mobility Controls |  |  | ✓ | ✓ |
| Testing Controls |  |  |  | ✓ |

\* $p < 0.1$ ; \*\* $p < 0.05$ ; \*\*\* $p < 0.01$
*Notes:* Results of panel-data regressions of the (log of) effective reproduction number ( $\mathcal{R}_t$ ) on indicator variables that are equal to 1 after the introduction of a non-pharmaceutical intervention (NPI) and 0 before the introduction. The sample consists of 14 European countries studied by Flaxman et al. \(2020\). Regressions always include country fixed effects; regressions in columns (2)–(4) also include days-since-outbreak fixed effects. Outbreak is defined as the date on which 100 cases of COVID-19 are reached. The regression with mobility controls in (3) includes the one- and two-week lags of the mobility index, constructed from Google \(2020\); see Appendix A.8 for details. The regression with testing controls in (4) controls for the change in the number of daily tests per capita conducted in the country. To allow for reasonably precise estimation of days-since-outbreak fixed effects, we only consider days after the outbreak for which we have data for at least 5 countries. Heteroskedasticity-robust standard errors in parentheses.

Column (1) of Table 2 provides estimated effects of NPIs when only country fixed effects are included. We observe a strong negative effect of lockdowns, social distancing, and measures of self isolation. Taken at face value, the estimates suggest that lockdowns reduce ℛ_*t*_ by 65%. School closures are not statistically significant in this specification. These regressions as well the point estimates are similar to the statistical analysis performed by Flaxman et al. (2020).

The regression with country fixed effects only, however, is likely misspecified. Implicitly, such a specification assumes that the only reason ℛ_*t*_ can fall is because of introduction of NPIs. However, ℛ_*t*_ would likely trend downwards even in the absence of any public policy interventions. First, ℛ_*t*_ tends to fall during an epidemic as the number of susceptibles is depleted. Second, people may adjust their behavior even in the absence of any policy measures. Failing to control for the dynamics of ℛ_*t*_ in the absence of NPIs therefore likely leads to an over-estimation of the effects of NPIs.

We acknowledge that obtaining credible counterfactuals in the present empirical context is extremely challenging. However, we can exploit the panel structure of the dataset to reduce the potential issues in the previous specification. We do so by including days-since-outbreak fixed effects. Intuitively, with such fixed effects we are comparing ℛ_*t*_’s in two countries (e.g., country A and country B) that are both five days from the outbreak (say), with a school closure in country A but not in country B.

The results from the regression with days-since-outbreak fixed effects are shown in column (2). The coefficient for lockdowns becomes substantially smaller in absolute value and less statistically significant. The coefficients for self-isolation and social-distancing measures are also reduced and lose some of their statistical significance. The coefficient for public events is highly statistically significant but positive rather than negative. A naïve interpretation would suggest that banning public events has a positive effect on ℛ_*t*_. More likely, however, is that the positive coefficient is due to countries where ℛ_*t*_ is declining more slowly being faster to ban public events. In the Appendix (Table A.4), we show that the results remain similar when the NPIs are included separately, reducing concerns about potential multicollinearity problems between the different NPI variables.

In column (3), we also include lagged mobility variables as additional controls. With mobility controls, the coefficient on lockdowns is further reduced. School closures and social-distancing measures are estimated to have a statistically-significant negative effect on ℛ_*t*_, with reducing ℛ_*t*_ by 18% and 11%, respectively.

A potential concern is that countries may introduce NPIs and simultaneously increase the number of tests for COVID-19 that they perform. To help alleviate this concern, in column (4) we add the change in the daily number of tests per capita as an additional explanatory variable. The data on daily tests per capita comes from *Our World in Data* (Our World in Data, 2020). With testing controls, most coefficients are no longer statistically significant. Note, however, that the sample size is reduced significantly as we do not have testing data for all countries in the sample.

We caution readers against over-interpreting the results of this section. Obtaining unbiased estimates of the true causal impact of NPIs is exceptionally challenging. As a result, even our best estimates might still suffer from statistical issues such as unobservable confounding variables or simultaneity bias. In particular, the timing of NPIs is not random. Countries that introduced NPIs earlier likely did so because they had previously observed a stubbornly high ℛ_*t*_. In that case, the dependent and independent variables would be simultaneously determined, yielding biased estimates. Moreover, since we cannot directly observe peoples’ attitudes towards COVID-19 or government policies, we cannot control for other variables affecting human behavior. These potential issues notwithstanding, we find that people adjusted their mobility patterns *before* the introduction of lockdowns. We believe that these findings bolster the importance of accounting for changes in human behavior when evaluating the effects of NPIs.

## 4 Conclusions and Limitations

In this paper we develop a new way to estimate the effective reproduction number of an infectious disease (ℛ). Our estimation method exploits a structural mapping between ℛ and the growth rate of the number of infected individuals derived from the basic SIR model. The new methodology is straightforward to apply in practice, and according to our simulation checks, it yields accurate estimates. We use the new method to track ℛ of COVID-19 around the world, and assess the effectiveness of public policy interventions in a sample of European countries.

The current paper faces several limitations. First, a local-level specification for the growth rate implicitly assumes that the growth rate of the number of infected individuals remains forever in flux. However, in the long-run, this growth rate must converge to zero. Since our model does not capture this feature, it seems likely that our estimated confidence bounds are overly conservative in the late stages of an epidemic. Second, when applying the model to cross-country data, one may achieve important gains in statistical efficiency if the model is estimated jointly for all countries (for example, by estimating a multivariate local-level model). Finally, for assessing the effects of NPIs more accurately, it would be desirable to collect data for a larger sample of countries.

Our estimates of ℛ for COVID-19 are based on a structural relationship derived from the SIR model. By using the SIR model, we omit some features of the disease that are likely important when modeling its spread. In particular, the SIR model abstracts away from incubation periods as well as transmission during the incubation period. Nevertheless, we prefer the SIR specification, for two reasons. First, in simulations, we find that our estimator produces accurate estimates even when the true model is SEIR rather than SIR (Appendix A.5). Second, we believe that the SIR model is likely to produce more reliable estimates in practice. To use the SEIR model, we would have to estimate the number of currently exposed individuals. Doing so would triple the number of model parameters. In particular, we would have to calibrate the (i) average duration of the incubation period (*κ*^−1^); and (ii) relative infectiousness of exposed and infectious individuals (*ϵ*); see Appendix A.5 for details. While *κ* is arguably constant across countries, *ϵ* is unlikely to be fixed over countries and over time. For example, greater mask usage is likely to reduce *ϵ* by differentially affecting transmission by symptomatic and pre- or asymptomatic individuals. Allowing for such time variation in *ϵ*, in addition to time-varying transmission rates (*β*_*t*_), is challenging. That said, it is possible to extend this paper’s ideas to models that are richer than the SIR model. Doing so may be an exciting avenue for future research.

Relative to existing methods for estimating ℛ, we combine basic epidemiological theory with standard time-series filtering techniques, particularly Kalman filtering. This approach leads to a transparent closed-form estimator. The simplicity of the estimator allows us to study some of its properties analytically (e.g., the effects of potential data problems). Differently from most existing approaches, our method can be applied using both Bayesian and frequentist techniques, and it does not require any tuning parameters beyond specifying the average serial interval. On the other hand, relative to less structural approaches such as that of Cori et al. (2013), our estimator may be more sensitive to potential model misspecification. Empirically, we find that our estimates and estimates obtained by the Cori et al. (2013) method are highly positively correlated (average correlation: 0.80). However, the correlations are not perfect, suggesting that there is value in combining both estimators when tracking infectious diseases. Hence, our methodology brings an additional instrument to the researcher’s toolbox.

In our empirical application, we find that lockdowns, measures of self-isolation, and social distancing all have statistically significant effects on reducing ℛ of COVID-19. However, we also demonstrate the importance of accounting for voluntary changes in behavior. In particular, most of the decline in mobility in our sample took place before lockdowns were introduced. This finding suggests that people respond to the risk of contracting the virus by changing their mobility patterns and reducing social interactions. Failing to account for such voluntary changes in behavior yields estimated effects of NPIs that are arguably too large.

Given that even our best estimates may still be biased, it is important to interpret these results cautiously. However, from an economic perspective, these findings point to large private incentives to avoid infection. These incentives can induce a contraction in economic activity as people voluntarily choose to self-isolate (Farboodi et al., 2020; Guerrieri et al., 2020; Krüger et al., 2020). As a result, even if countries lift the NPIs that are currently in place, it is not clear whether people would voluntarily return to their pre-pandemic mobility and consumption patterns. Our real-time estimator may be used to track the dynamics of COVID-19 as the current restrictions are relaxed.

## Data Availability

All the data used for the paper is available in our website.

http://trackingr-env.eba-9muars8y.us-east-2.elasticbeanstalk.com/

## Appendix A Supplementary Methods and Materials

### A.1 SIS Model

We now show that the estimator in Eq. (3) can also be obtained when the dynamics of the disease follow the SIS model. The SIS model, again in discrete time, is given by

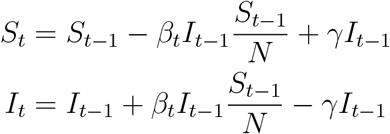

The only difference from the SIR model in Eq. (1) is that formerly infected individuals do not obtain immunity after recovery and instead again join the pool of susceptibles. As is well known, the basic reproduction number in the SIS model is the same as in the SIR model (e.g., Chowell and Brauer, 2009) and given by 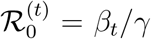. Since the law of motion for *I*_*t*_ in the SIS model is the same as in the SIR model, we can repeat the same steps as in the benchmark analysis to arrive at Eq. (3).

### A.2 Generalized SIR Model

In this section, we show that we can also obtain the estimator in Eq. (3) from a generalized version of the SIR model with stochastic shocks. Specifically, we consider the following generalized SIR model:

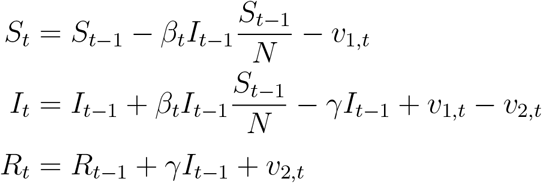

Differently from the baseline model, we introduce random shocks *v*_1,*t*_ and *v*_2,*t*_. The shocks are i.i.d., and the time-varying support of *v*_1,*t*_ is [0, *S*_*t*−1_ − *β*_*t*_*I*_*t*−1_/*N*], while the support of *v*_2,*t*_ is [0, *I*_*t*−1_ + *β*_*t*_*I*_*t*−1_*S*_*t*−1_/*N*]. We also assume that 𝔼_*t*−1_[*v*_1,*t*_ − *v*_2,*t*_] = 0, so that the conditional expectation 𝔼_*t*−1_[*I*_*t*_] coincides with the value for *I*_*t*_ given by the noiseless SIR model. With these modifications, the model can capture rich patterns of infectious disease dynamics. For example, “super spreader events” can be modeled either as *v*_1,*t*_ shocks or as a spike in *β*_*t*_. The model can also capture richer forms of population structures than the baseline SIR model. For example, if individuals who are more infectious (e.g., those with more connections in a network model) are more likely to become infected first, that can be captured by assuming that *β*_*t*_ becomes lower over time.

Defining the time-varying basic reproduction number as 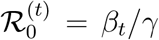, and 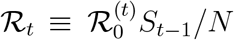, we obtain that

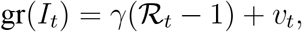

where *v*_*t*_ ≡ (*v*_1,*t*_− *v*_2,*t*_)/*I*_*t*−1_. Taking expectations on both sides of the equation, we arrive at

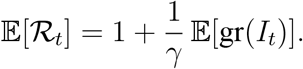

Hence, the generalized SIR model of the present section leads to the same estimator as the baseline SIR model in Eq. (1).

Finally, we note that if *γ* varies deterministically over time, the equation above remains essentially unchanged, the only difference being that *γ* is replaced by *γ*_*t*_. If *γ*_*t*_ follows a non-degenerate stochastic process, then the estimator for 𝔼[ℛ_*t*_] would need to correct for the covariance between *γ*_*t*_ and ℛ_*t*_.

### A.3 Foundation for the Local-Level Model

When estimating ℛ_*t*_, we use a local-level specification for the growth rate of the number of infected individuals. In this section, we show that the local-level model arises naturally in the SIR model in the early stages of an epidemic when the transmission rate follows a random walk.

Specifically, consider the generalized SIR model in Appendix A.2. We now specialize the process for the transmission rate *β*_*t*_ to be a random walk:

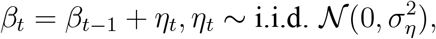

with a given initial value *β*_0_ > 0. We calculate that

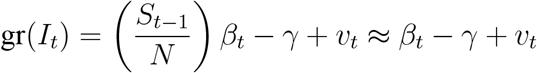

in the early stages of the epidemic when *S*_*t*_ ≈ *N*. Defining the effective reproduction number early on in the epidemic as ℛ_*t*_ = *β*_*t*_/*γ*, we therefore have directly that the growth rate of *I*_*t*_ follows a local-level model with

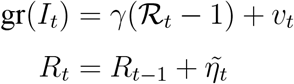

where 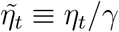 Provided that the distribution of *v*_*t*_ can be approximated with a normal distribution, we directly obtain the specification in Eq. (5). Alternatively, to obtain an exact normal local-level model, we could assume that *v*_1,*t*_ = *v*_2,*t*_ = 0 (no shocks in the original model, just as in Eq. (1)) but that instead of observing the true growth rate gr(*I*_*t*_), we only observe gr(*I*_*t*_) + *ε*_*t*_ where *ε*_*t*_ is i.i.d. normally distributed mean-zero measurement error.

### A.4 Gibbs-Sampling Algorithm

In this section we discuss how the parameters of the state-space model in Eq. (5) can be estimated with a Gibbs-sampling algorithm *à la* Carter and Kohn (1994). Besides being a natural robustness check to our methodology, this algorithm uses a different approach to ensure the non-negativity of ℛ_*t*_.

To use the Kalman filter, we need to estimate 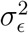 and 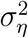 For the Gibbs sampler, we break the model down into conditional densities from which we can sample iteratively. The algorithm is the following:

1. Conditional on 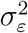 and 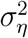, use the Kalman filter to infer the state vector ℛ_*t*_;
2. Conditional on the sequence of ℛ_*t*_ computed in the previous step, take samples of 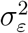 and 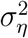 from their prior distributions;
3. Conditional on the new draws of 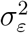 and 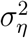, estimate ℛ_*t*_;
4. Verify that each element of ℛ_*t*_ is positive. If yes, store the draws of 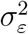 and 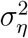. If not, discard the draws and repeat step 2;
5. Compute the Kalman smoother;
6. Iterate forward for as many replications as needed.

Finally, we contrast the estimates of ℛ_*t*_ obtained with the Gibbs sampler to our baseline estimates. We obtain a correlation of 0.85 between the two sets of estimates. Credible intervals are highly correlated as well. Overall, we conclude that our estimates are similar across different statistical estimation approaches.

### A.5 SEIR Model: Monte Carlo Simulation

Our estimation method uses a structural mapping between ℛ_*t*_ and gr(*I*_*t*_) derived from the basic SIR model. While we can generalize the baseline SIR model to include stochastic shocks (Appendix A.2), and the estimator remains valid when the disease follows an SIS model (Appendix A.1), the model is nevertheless restrictive. In particular, it ignores incubation periods as well as transmission during the incubation period. These features are likely especially important when modeling COVID-19.

We now perform a simulation exercise to see how our estimator of ℛ_*t*_ performs in a richer model that accounts for these additional features. Specifically, we consider an SEIR model in which the exposed are infectious:

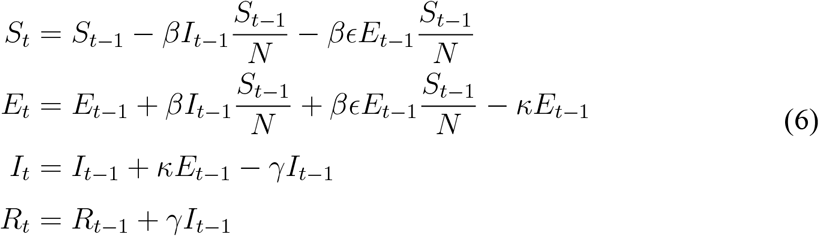

Here, *E*_*t*_ denotes the number of individuals that are exposed at day *t, κ* is the daily transition rate from exposed to infected, and *ϵ* ∈ [0, 1] measures the degree to which the exposed are less infectious than the infected. If *ϵ* = 0, the exposed are not infectious at all, and we obtain the benchmark SEIR model. If *ϵ* = 1, the exposed are as infectious as the infected, and the model is isomorphic to the standard SIR model.

We calibrate the parameters following Wang et al. (2020) who apply the benchmark SEIR model (with *ϵ* = 0) to study the dynamics of COVID-19 in Wuhan. In particular, we use *κ* = 1/5.2 and *γ* = 1/18 as in Wang et al. (2020). Then, we set *ϵ* = 2/3, following Ferguson et al. (2020) who assume that symptomatic individuals are 50% more infectious than the asymptomatic (that is, *ϵ*^−1^ = 1.5). Finally, we choose *β* by targeting a basic reproduction number of ℛ_0_ = 2.6, again as in Wang et al. (2020). In the model above, ℛ_0_ is given by ℛ_0_ = *β*/*γ* + *βϵ*/*κ*, implying *β* = ℛ_0_*γκ*/(*γϵ* + *κ*). The formula yields *β* ≈ 0.12. Finally, we set *S*_0_ = 11 × 10^6^ (approximating the population size of Wuhan), *E*_0_ = *R*_0_ = 0, and *I*_0_ = 1.

The Monte Carlo design is as follows. First, we simulate the deterministic system in Eq. (6) using the parameters above. Then, we calculate the growth rate in the true number of infected individuals, i.e., gr(*I*_*t*_) = *I*_*t*_/*I*_*t*−1_ − 1. However, instead of knowing the true growth rate, the statistician is assumed to observe a noisy version of it given by 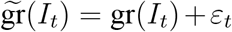 Here, *ε* is an i.i.d. normal disturbance with mean zero and standard deviation of 0.10. The standard deviation of the disturbances is roughly equal to the range of the true growth rates. Hence, the amount of noise used in the simulation is fairly large. For each realization of the disturbances, we estimate ℛ_*t*_ using our method. As in our empirical application, only data after 100 total cases have been reached is used.

We investigate two values for *γ*_est._ that are used when estimating ℛ_*t*_ via Eq. (3). First, we consider a situation in which the statistician uses the correct time that individuals are infected, given by *γ*_est._ = (*γ*^−1^ + *κ*^−1^)^−1^ where *γ* and *κ* are the true parameter values of the SEIR model. Second, we investigate a case in which the statistician incorrectly thinks that individuals are infectious only for ten days (*γ*_est._ = 1/10). We repeat the process for 10,000 Monte Carlo replications.

**Figure A.1.**
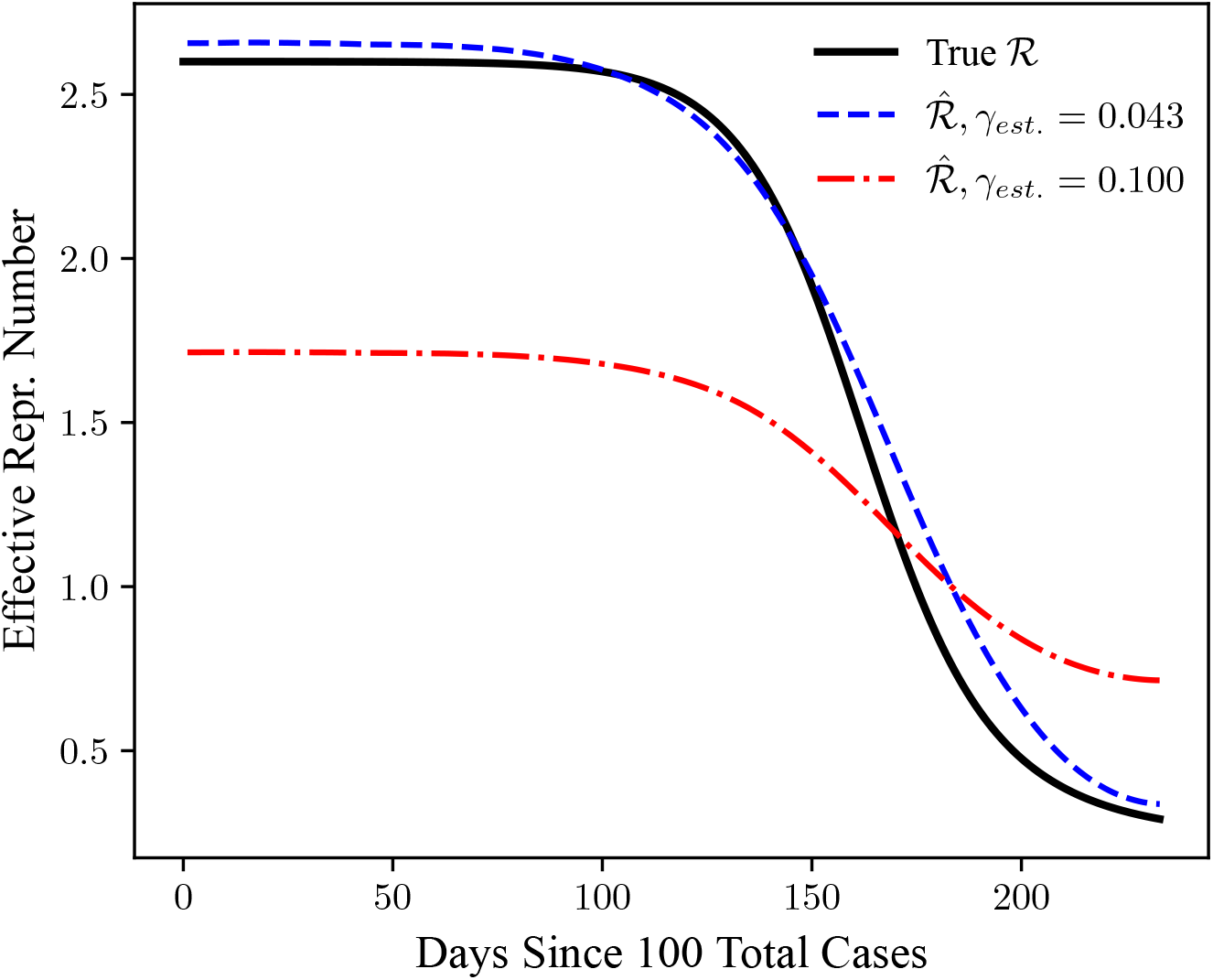
Monte Carlo Simulation: Effects of Misspecification. *Notes*: Estimates of the effective reproduction number (ℛ_*t*_) when the true dynamics of the disease follow an SEIR model. We investigate two values for *γ*_est._, the transition rate from infected to recovered, that are used when estimating ℛ_*t*_. First, we use the correct value of *γ*_est._ = (*γ*^−1^ + *κ*^−1^)^−1^ ≈ 0.043. Second, we use a misspecified values of *γ*_est._ = 1/10. Average values from 10,000 Monte Carlo replications are shown. See text for more details.

The results of the Monte Carlo simulation are shown in Figure A.1. When the statistician uses the correct number of days that an individual is infectious (that is, taking into account the incubation time), the estimates of ℛ_*t*_ from our method are very close to their true theoretical values. That is in spite of the fact that our estimator for ℛ_*t*_ is derived assuming that the dynamics of the disease are described by an SIR model. However, we also show that if the statistician misspecifies the number of days than an individual is infectious (assuming 10 days instead of the true number of 23.2 days), the estimates of ℛ_*t*_ are substantially biased, especially in the early stages of the epidemic. As is to be expected from Eq. (3), underestimating the number of days that an individual is infectious leads to a downwards bias in the estimates of ℛ_*t*_ early on in the epidemic (when ℛ_*t*_ > 1), and upwards bias when the true ℛ_*t*_ falls below one. Overall, the results imply that the new method performs well when estimating ℛ_*t*_ even when the true dynamics of the disease do not follow the SIR model, provided that the duration of infectiousness used in the estimation is sufficiently accurate.

### A.6 Effects of Potential Data Issues

We now discuss the effects of various data issues on the performance of our estimator.

#### Reporting delays

In practice, data may be subject to significant reporting delays. For example, suppose that due to testing constraints there is a lag of *ℓ* days between the date that an individual becomes infected and the date on which the case is registered. In this case, the estimates of ℛ_*t*_ would also be subject to delay of *ℓ* days. If there are significant reporting delays, one may first obtain, say, one-week-ahead forecasts of new cases, and then use these forecasts to construct a time series for *I*_*t*_.

#### Imperfect detection

A natural worry with any estimator of ℛ_*t*_ is that it may be substantially biased if not all of infected individuals are detected. Given the simplicity of our estimator, we can analytically assess the effects of imperfect detection.

Suppose that the true numbers of susceptible, infected, and recovered individuals are given by 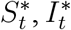, and ℛ_*t*_ respectively. Their evolution is the same as in Eq. (1). However, we only observe 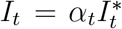, where 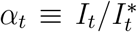 is the *detection rate*. In practice, *α* is typically less than one, although the mathematical calculation below does not require this.

With this notation,we have that

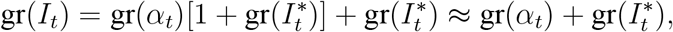

since 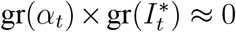 at a daily frequency; the approximation is exact in continuous time. Using the approximation above and Eq. (2), we therefore obtain that the bias of the estimator under imperfect detection is given by

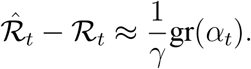

We now discuss several cases of practical importance:

- Constant detection rate (*α*_*t*_ = *α*). If the detection rate is constant over time, then our estimator is unbiased, and 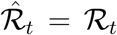 Hence, for example, even if we only detect 10% of the infectives (but the fraction detected remains constant over time), the estimator remains unbiased. Note that if the number of tests increases over time, that is *not* inconsistent with *α*_*t*_ = *α* given that the number of infected individuals is likely to be growing at the same time.
- Constant growth in the detection rate (gr(*α*_*t*_) = *g*_*α*_). If the growth rate of *α*_*t*_ is constant over time, then our estimate of ℛ_*t*_ is biased upwards if *g*_*α*_ > 0 and downwards if *g*_*α*_ < 0. Note, however, that we are often mostly interested in the *trend* of ℛ_*t*_ over time and whether the trend is affected by various policy interventions. The trend in ℛ_*t*_ is estimated accurately even if *g*_*α*_*̸*= 0. Intuitively, constant growth in the detection rate leads to a level bias, but the slope is still estimated correctly.
- Detection rate converges over time (*α*_*t*_ → *α*). The final case of interest occurs when the detection rate converges to a constant over time. For example, if everyone is detected towards the end of the epidemic, we would have *α*_*t*_ → 1. Since our method uses Kalman-filtering techniques to estimate the growth rate of *I*_*t*_, transient fluctuations in *α*_*t*_ would have a limited effect on the estimates of ℛ_*t*_ later on in the sample. Given that we are often precisely interested in the behavior of ℛ_*t*_ in the later stages of the epidemic (when the detection rate is likely fairly constant), our method would still yield reliable estimates.

To provide a quantitative illustration, we perform a small-scale Monte Carlo study. We choose the parameters of the simulation to match our empirical estimates. First, we use our empirical estimates of ℛ for the world as a whole for the first 50 days of the sample as the true values of ℛ. From these values of ℛ, we calculate the true (but unobserved) values of the growth rate of the number of infected individuals as 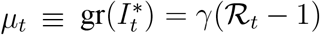, using *γ* = 1/7. Finally, the observed growth rate—as seen by the statistician—is generated as gr(*I*_*t*_) = gr(*α*_*t*_)(1 + *µ*_*t*_) + *µ*_*t*_ + *ε*_*t*_. We use the empirical estimate of 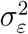 to simulate *ε*_*t*_ shocks (i.i.d. draws from a normal distribution with mean zero). We then apply our estimator to the generated data on gr(*I*_*t*_), using the same value of *γ* = 1/7 to back out ℛ.

In the simulation we consider three underdetection scenarios:

- *Constant underdetection*. Given our analytical results, it is immaterial what percentage of cases is detected, as long as that percentage is constant over time.
- *Ramp-up in testing*. Next, we consider a situation in which—due to an increase in the number of tests performed—the fraction of detected infected individuals goes up from *α*_0_ = 0.10 to *α*_14_ = 0.15 (an increase of 50%) over a period of two weeks. After the two weeks, the detection probability remains constant at 0.15. Again, the precise values of *α*_0_ and *α*_14_ are irrelevant, and what matters is the growth rate in the detection rate.
- *Stochastic underdetection*. Finally, we suppose that the detection rate satisfies gr(*α*_*t*_) = *ϕ* gr(*α*_*t*−1_) + *ν*_*t*_ where *ν*_*t*_ is an i.i.d. normally distributed shock. Intuitively, the detection rate is assumed to be unconditionally constant, but its growth rate is stochastic and follows an AR(1) process. We set the persistence parameter to *ϕ* = 0.75 in order to allow for fairly long-lasting deviations from the average detection rate. To ensure that the variance of gr(*I*_*t*_) remains constant across simulations (to ensure an apples-to-apples comparison), we suppose that 50% of the noise in the observed growth rate comes from variation in *α*_*t*_, with the other 50% coming from the *ε*_*t*_ shocks. (Here, by “noise” we refer to the variation in gr(*I*_*t*_) that is not solely due to the variation in *µ*_*t*_, namely, gr(*α*_*t*_)(1 + *µ*_*t*_) + *ε*_*t*_.) Denoting our estimates of the variance of the growth rate by 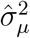 and the variance of the irregular component by 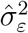, we therefore set 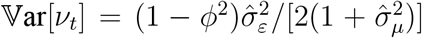 and 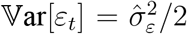. We draw the initial value for gr(*α*_0_) from its unconditional distribution.

**Figure A.2.**
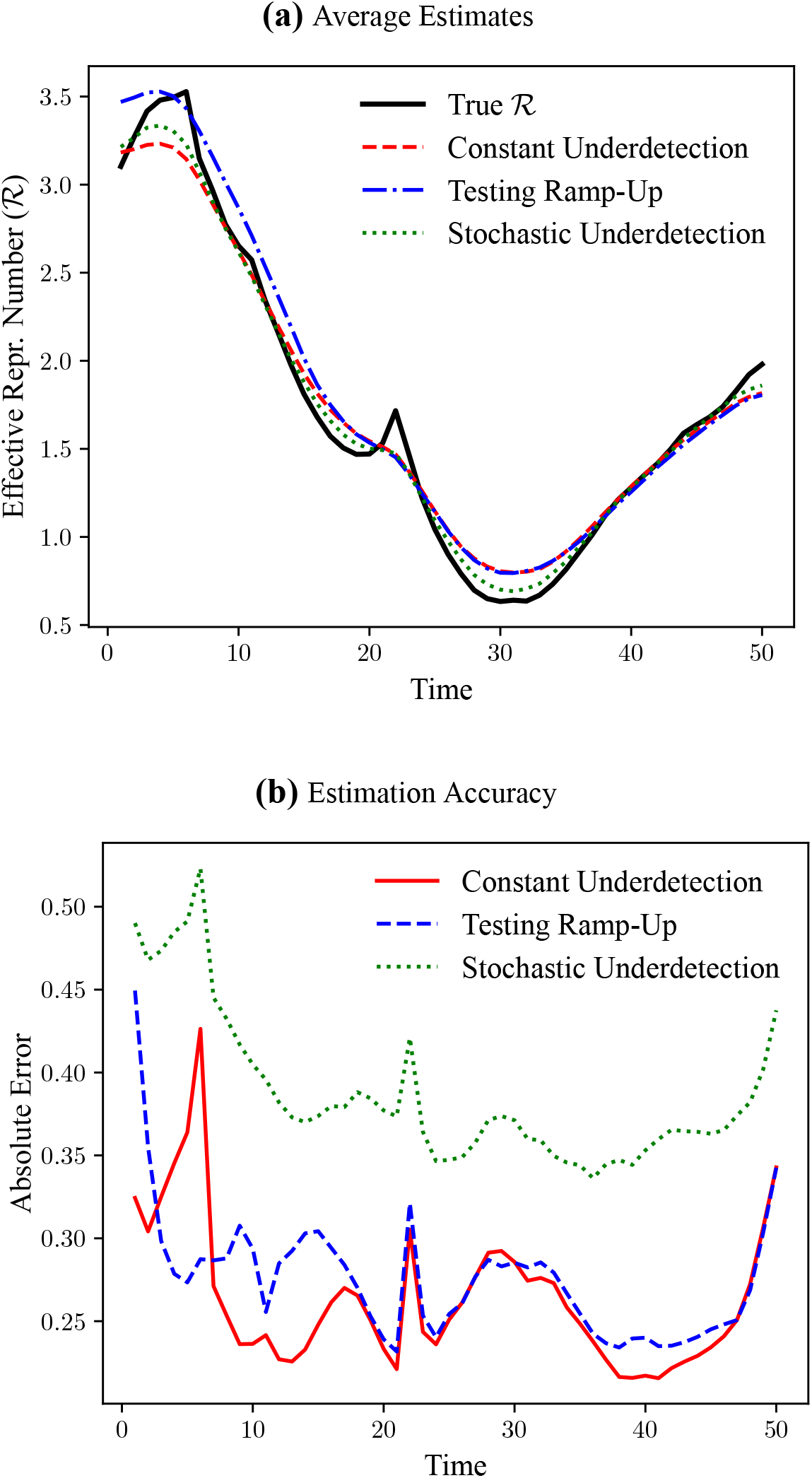
Monte Carlo Simulation: Effects of Underdetection. *Notes*: Estimates of the effective reproduction number (ℛ_*t*_) when the true number of infected individuals is measured with error. The true values for ℛ_*t*_ are given by our empirical estimates of ℛ_*t*_ for the world as a whole in the first 50 days of the sample, and the standard deviation of the irregular component is chosen to match our empirical estimates. *Constant Underdetection*: A constant fraction of infected individuals is detected. *Testing Ramp-Up*: The fraction of detected individuals increases by 50% in the first two weeks of the sample. *Stochastic Underdetection*: The fraction of detected individuals is stochastic, and its growth rate follows an AR(1) process. Average values from 1,000 Monte Carlo replications are shown. *Top panel*: Average estimates. *Bottom panel*: average absolute error of the estimates.

The results of the Monte Carlo simulation are summarized in Figure A.2. As seen in the top panel, the estimated average effective reproduction numbers are fairly close to their theoretical values in all three scenarios. In the *Testing Ramp-Up* scenario, the estimates are biased upwards at the beginning of the sample, but the amount of bias is quantitatively relatively small. In addition, the estimates converge to those in the other two scenarios quite quickly. In all three scenarios, the estimates are able to pick up the reversal in the trend of ℛ that happens at around time 30. The bottom panel of the figure plots the average absolute error of the estimates. As expected, the estimates are least accurate in the *Stochastic Undertesting* case, and mostly within 0.25–0.30 of the true ℛ in the *Constant Underdetection* and *Testing Ramp-Up* scenarios.

Finally, in Figure A.3 we provide Monte Carlo estimates of the coverage frequency of credible intervals obtained by our estimation procedure. As seen in the graph, the credible intervals in the *Constant Undertesting* and *Testing Ramp-Up* scenarios have good coverage properties, with conservative confidence bounds. The credible bounds are narrower in the *Stochastic Underdetection* scenario, resulting in lower than nominal coverage frequency. However, the size distortion appears not too severe, especially considering the small sample size.

#### Imported cases

Our estimates may be biased if the fraction of cases that is imported changes over time (the previous results on imperfect detection apply to misclassification because of imported cases, too). If the source of infections is known, it is possible to correct for the issue by simply not including imported cases when constructing the time series for *I*_*t*_.

**Figure A.3.**
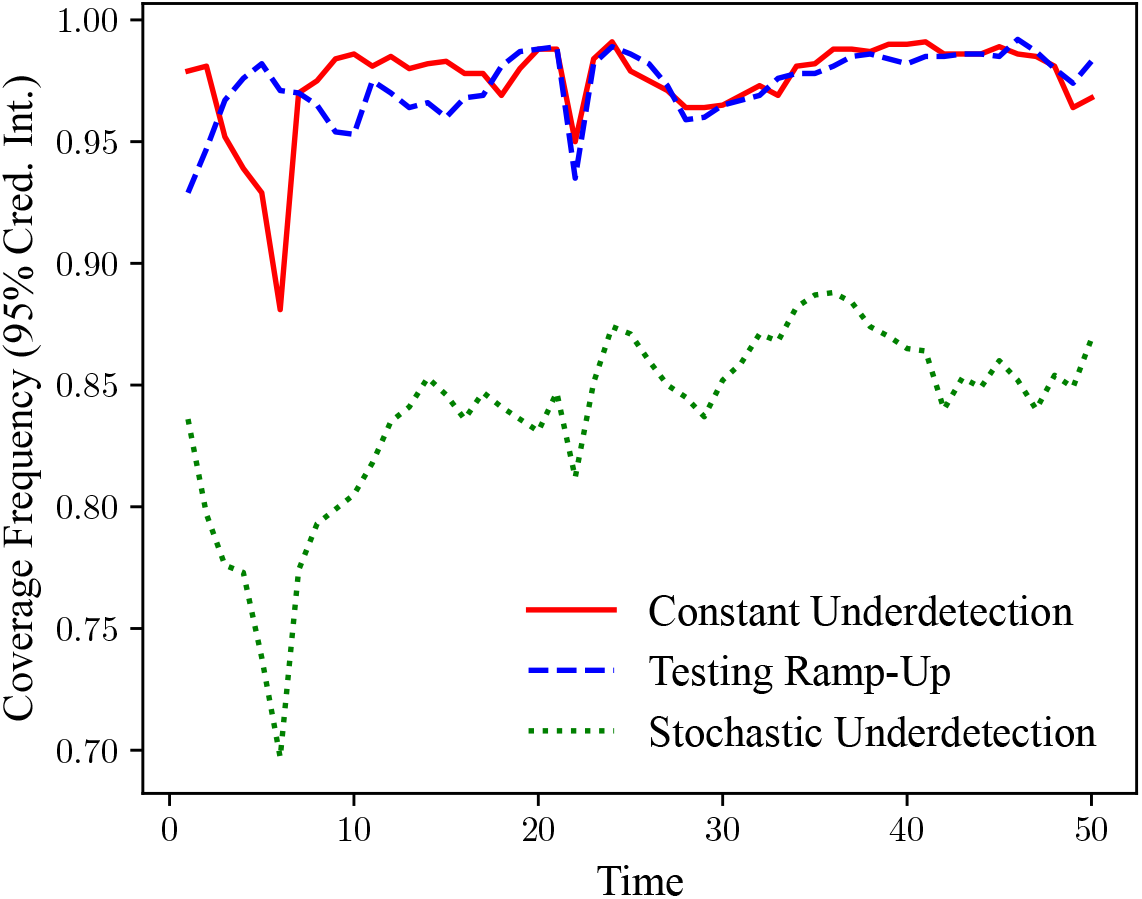
Monte Carlo Simulation: Coverage Frequency. *Notes*: Coverage frequency of 95% credible intervals: Monte Carlo results. We investigate three scenarios for measurement error. *Constant Underdetection*: A constant fraction of infected individuals is detected. *Testing Ramp-Up*: The fraction of detected individuals increases by 50% in the first two weeks of the sample. *Stochastic Underdetection*: The fraction of detected individuals is stochastic, and its growth rate follows an AR(1) process. Average values from 1,000 Monte Carlo replications are shown. See text and Figure A.2 for more details.

**Table A.1.** Priors.

| Parameter | Prior |
| --- | --- |
| Precision of irregular component ($1/\sigma_\varepsilon^2$) | Gamma(0.17985, 0.00099) |
| Signal-to-noise ratio ($q \equiv \sigma_\eta^2/\sigma_\varepsilon^2$) | Gamma(0.07781, 0.54261) |
| Initial value ($\mu_0$) | $\mathcal{N}(0.35, 0.5^2)$ |

*Notes:* Priors used in the Bayesian estimation of $\mathcal{R}_t$ . See text for description on how the priors for the precision of the irregular component ( $1/\sigma_\varepsilon^2$ ) and the signal-to-noise ratio ( $q \equiv \sigma_\eta^2/\sigma_\varepsilon^2$ ) are calibrated based on cross-country frequentist estimates.

### A.7 Estimation Details

To estimate ℛ_*t*_ of COVID-19, we use Bayesian filtering methods. We employ the following strategy to calibrate the prior distributions. First, we estimate a local-level model for gr(*I*_*t*_) using a frequentist Kalman filter with diffuse initial conditions. In particular, we estimate the following model:

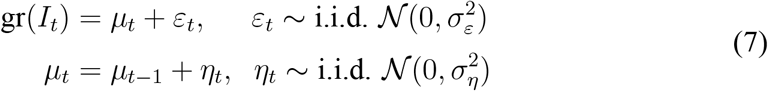

The model is the same as in Eq. (5), except for a slight simplification in notation.

The procedure yields maximum likelihood estimates of 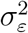 (variance of the irregular component) and the signal-to-noise ratio 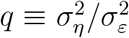 for each country in the sample (with 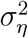 denoting the variance of the level component). We then use the distribution of 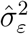 and 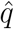 across countries to calibrate the priors for the precision of the irregular component 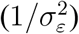 and the signal-to-noise ratio (*q*). To ensure that the priors are not too “dogmatic,” we inflate the variance of the estimates by a factor of 3 when calibrating the prior distributions. We use a gamma prior for both the signal-to-noise ratio and the precision of the irregular component, and we calibrate the parameters of the gamma distribution by matching the expected value and variance of the gamma-distributed random variables to their sample counterparts. Finally, we use a fairly uninformative normal prior for the initial value of the smoothed growth rate. The resulting priors are given in Table A.1.

Intuitively, these priors shrink the estimates of the precision and signal-to-noise ratio for each country towards their grand mean (average across countries). Such Bayesian shrinkage ensures that the parameter estimates are well behaved even though the sample size for many countries is fairly small, and the data are often noisy. We use the Stan programming language (Gelman, Lee and Guo, 2015) to specify and estimate the Bayesian model. In particular, we use the `pystan` interface to call Stan from Python.

### A.8 Empirical Validation

In this section, we perform two empirical validation exercises to check the performance of our estimates in practice.

Since our estimates are based on data on new cases, they may be misleading if new cases are subject to significant measurement problems. To help assuage this concern, we now perform the following exercise. We ask whether *current* values of ℛ_*t*_ help predict *future* growth in deaths. Since deaths are likely to be measured more accurately, this exercise provides a test of whether our estimates contain meaningful information and are not contaminated by data problems.

Formally, we consider the following regression:

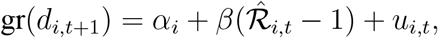

where *i* denotes a particular country, and *t* indexes calendar weeks. Although our original data is daily, we aggregate to a weekly frequency; otherwise, measures of the growth rate of new deaths are too noisy. In addition, we only include weeks after the cumulative number of COVID-19 deaths has reached 50.After imposing these sample restrictions, we are left with 270 country-week observations across 68 countries. Given that we have panel data, we can include country fixed effects *α*_*i*_ to account for time-invariant unobserved heterogeneity (such as differences in average age—a key correlate of COVID-19 mortality (Verity et al., 2020)—or family structures). The relationship given above is predicted by the baseline SIR model. Specifically, consider Eq. (2). Letting CFR = *d*_*t*_/*I*_*t*−*ℓ*_ denote the case fatality rate (assumed to be constant over time), with *ℓ* standing for the average time between becoming infected and death, we have that gr(*d*_*t*_) = gr(*I*_*t*−*ℓ*_), yielding the regression equation above.

**Figure A.4.**
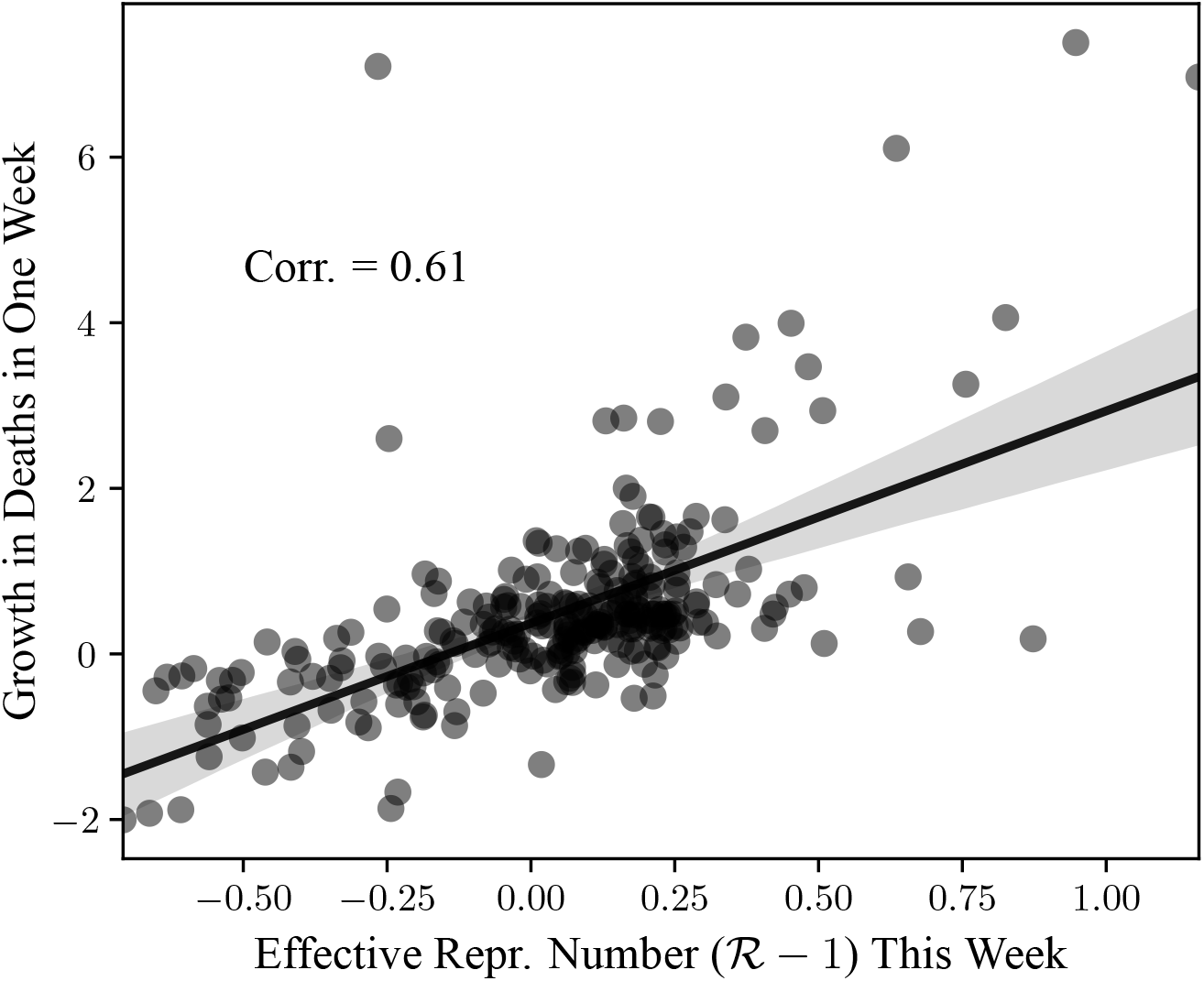
ℛ_*t*_ and Future Deaths. *Notes*: Relationship between current estimates of the effective reproduction number (ℛ_*t*_) and the growth rate of the number of new deaths in one week. The data is aggregated to a weekly frequency. Both variables are residualized to subtract country fixed effects by performing the within transformation. Only data after the cumulative number of deaths reaches 50 is included in the scatter plot. We include all countries in the John Hopkins database for which we have at least 20 observations after the outbreak. We remove data for the week of 2020-04-13–2020-04-19 in China that contain a large number of deaths that were previously unrecognized.

**Figure A.5.**
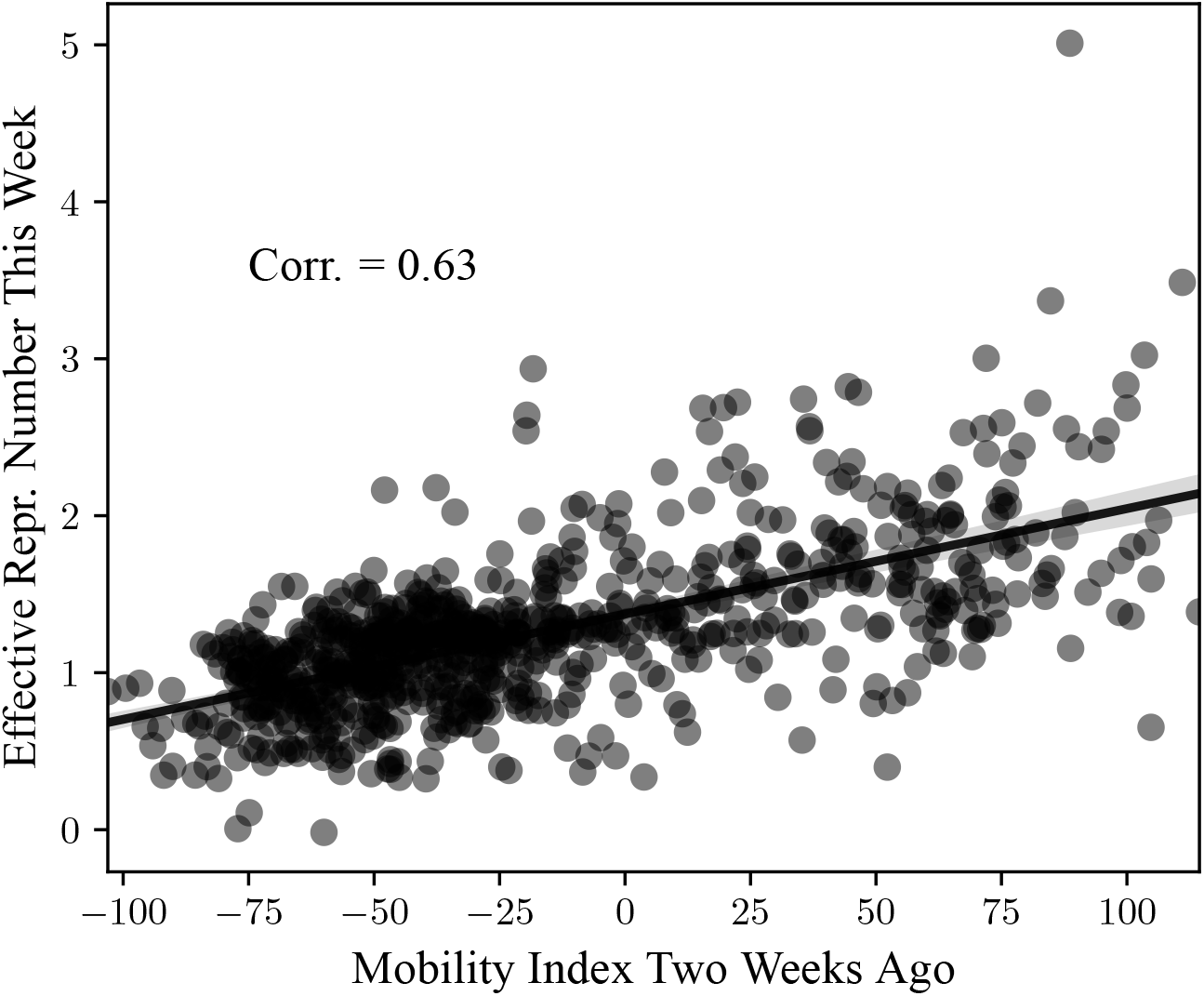
ℛ_*t*_ and Past Mobility. *Notes*: Relationship between current estimates of the effective reproduction number (ℛ_*t*_) and value of the movement index two weeks ago (first principal component of the six movement categories in Google (2020)). The data is aggregated to a weekly frequency. Both variables are residualized to subtract country fixed effects by performing the within transformation. We include all countries in the John Hopkins database for which we have at least 20 observations after the outbreak.

The relationship is shown in Figure A.4. In the scatter plot, both variables are residualized to remove country fixed effects. We observe a strong positive relationship between the value of ℛ_*t*_ this week and the growth in deaths one week later (corr. = 0.61). In Supplementary Figure (Figure A.9), we demonstrate that there is also positive correlation (corr. = 0.48) between ℛ_*t*_ and deaths two weeks later. We note that while the average medical duration from the onset of symptoms to death for COVID-19 is longer than two weeks (around 18 days, see Verity et al., 2020), the duration from *reported* cases to deaths is likely to substantially shorter because of reporting delays. For example, Hortaçsu, Liu and Schwieg (2020) assume that new cases of COVID-19 are reported with a lag of 8 days in their baseline calculations (5 days for symptoms to appear, consistent with the evidence from Lauer et al. (2020) and Park et al. (2020), as people are unlikely to be tested without exhibiting symptoms, and an additional 3 days to capture delays in obtaining test results, based on andecdotal reports from the US). Since deaths are likely reported in a timely manner, if new cases are reported with a lag of 8 days, we would expect an average duration of around 10 days (≈1.43 weeks) between reported cases and reported deaths.

As a second validation check, we ask whether our estimates of ℛ_*t*_ are correlated with past movement data, as it should be if the estimates are meaningful. For information on movement, we use aggregated smartphone location data collected by Google and published in their “COVID-19 Community Mobility Reports” (Google, 2020). Google provides data on percentage changes in movement for six types of places: (i) groceries and pharmacies; (ii) parks; (iii) transit stations; (iv) retail and recreation; (v) residential; and (vi) workplaces. Since the six categories are strongly correlated, we take the first principal component of the six categories (the first principal component explains 83.03% of the total variance in the data). We refer to the first principal component as the “Mobility Index.” As before, we only consider weeks after the cumulative number of confirmed COVID-19 cases in the country reaches 100. After imposing these restrictions, we are left with 792 country-week observations over 100 countries. Note that the number of countries is less than 124 because Google does not provide mobility data for all countries in our original sample. As shown in Figure A.5, current estimates of ℛ_*t*_ are strongly correlated with the value of the mobility index two weeks ago (corr. = 0.63).

For both validation exercises performed in the present section, we include all countries for which we have at least 20 observations after the onset of the epidemic (100 cumulative cases of COVID-19 reached) and satisfy any additional sample restrictions, as outlined above. If we narrow the sample down to countries with more and higher-quality data—such as the sample of European countries analyzed in Section 3.3—the correlations generally become substantially stronger. Hence, we consider the tests of the present section to be conservative.

### A.9 Power Analysis

In this section, we study the statistical power of the empirical analysis in Section 3.3 using a Monte Carlo simulation.

We now describe the design of the power study. Intuitively, we simulate data using a stochastic process that is calibrated to match the properties of the observed data. We then simulate a sharp drop in the effective reproduction number—say, because of a lockdown. We apply our estimator to the simulated data and ask how often this abrupt change is detected by the estimation procedure.

**Table A.2.**
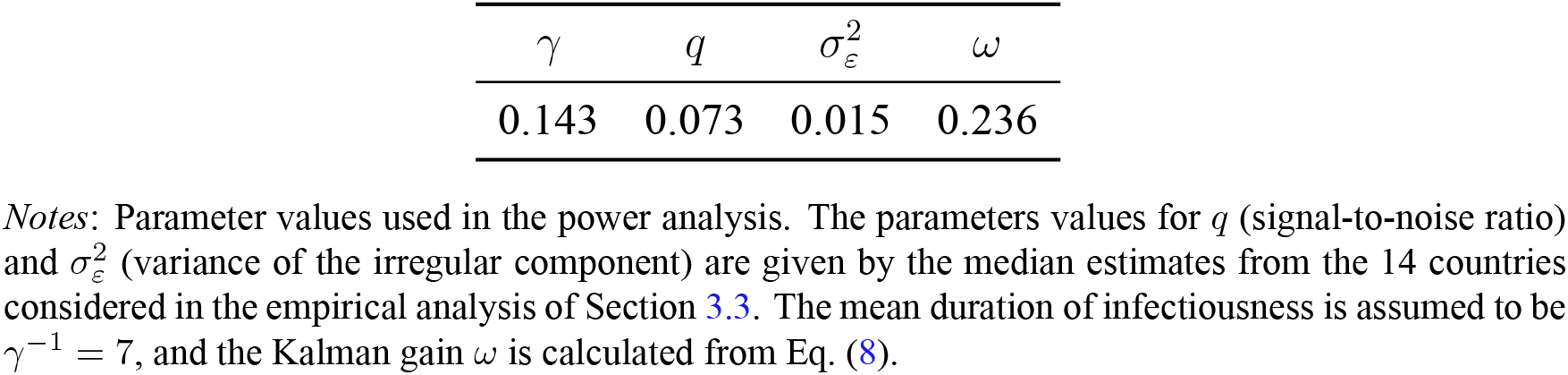
Power Analysis: Parameter Values.

| $\gamma$ | $q$ | $\sigma_\varepsilon^2$ | $\omega$ |
| --- | --- | --- | --- |
| 0.143 | 0.073 | 0.015 | 0.236 |

*Notes:* Parameter values used in the power analysis. The parameters values for $q$ (signal-to-noise ratio) and $\sigma_\varepsilon^2$ (variance of the irregular component) are given by the median estimates from the 14 countries considered in the empirical analysis of Section 3.3. The mean duration of infectiousness is assumed to be $\gamma^{-1} = 7$ , and the Kalman gain $\omega$ is calculated from Eq. (8).

To simplify the notation, we use the parametrization in Eq. (7). Optimal nowcasts from the local-level model in the steady state can be written as (Muth, 1960; Shephard, 2015, Section 3.4):

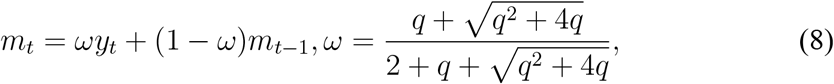

where we denote 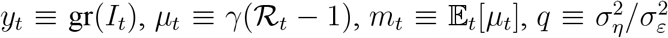 is the signal-to-noise ratio, and *ω* is the steady-state Kalman gain. Hence, nowcast errors, *m*_*t*_ − *µ*_*t*_, follow an AR(1) process with

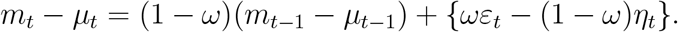

Given that the shocks *ε*_*t*_ and *η*_*t*_ are uncorrelated, the variance of nowcast errors is

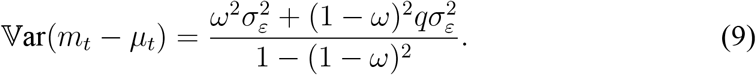

The design of the power analysis is as follows:

1. We set *γ* = 1/7, and calibrate the remaining parameters of the data-generating process (*q* and 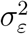) using the median values of the empirical estimates from Section 3.3. The resulting parameter values are given in Table A.2.
2. We simulate *µ*_*t*_ = *γ*(ℛ_*t*_ − 1). We initially set *µ*_0_ = 1/7, implying an effective reproduction number of 2. At time 1, we simulate an abrupt decline in ℛ_*t*_ by setting *µ*_1_ = 1/14, yielding a new effective reproduction number of 1.5, or a decline of 25%. For 2 ≤ *t* ≤ 14, we simulate *µ*_*t*_ as a random walk, as in Eq. (7).
3. We simulate the observed growth rate of the number of infected individuals, *y*_*t*_ = gr(*I*_*t*_), as *y*_*t*_ = *µ*_*t*_ + *ε*_*t*_ where *ε*_*t*_ is an i.i.d. normal random variable with mean zero and variance 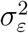.
4. We simulate the nowcasts *m*_*t*_. We draw the initial nowcast *m*_0_ from a normal distribution with mean 1/7 and variance given in Eq. (9), and simulate further
5. We repeat steps 2–4 for 14 times, to simulate data for 14 “countries”, as in the empirical application, and obtain estimates of the effective reproduction number by averaging across the 14 “countries”.
6. We repeat steps 2–5 for 10,000 Monte Carlo replications.

**Figure A.6.**
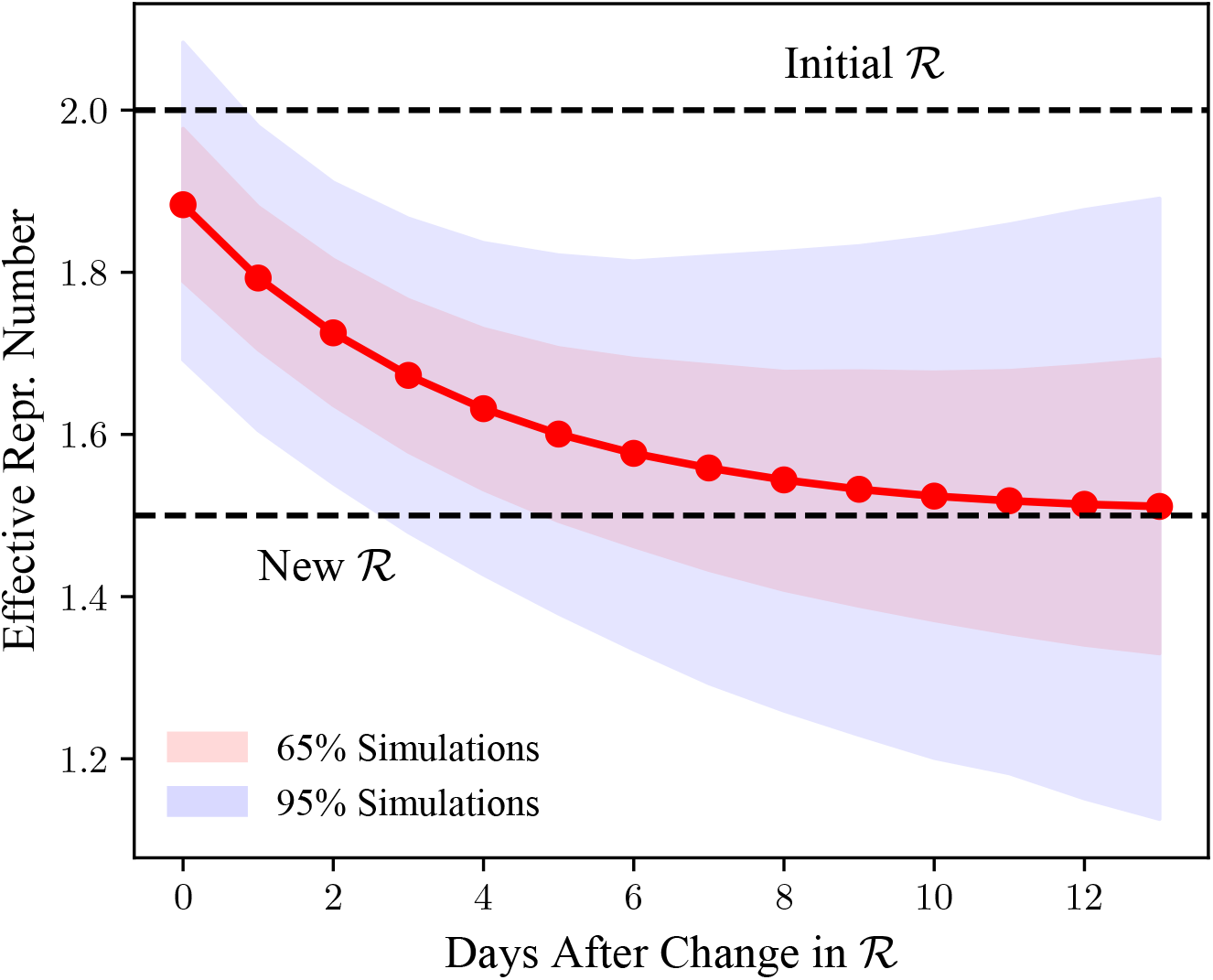
Power Analysis: Monte Carlo Results. *Notes*: Power study of the statistical analysis of Section 3.3 (effects of non-pharmaceutical interventions on ℛ_*t*_, the effective reproduction number). We simulate an abrupt change in _*t*_ from 2.0 to 1.5 using a data-generating process that is calibrated to match our empirical estimates in Section 3.3. We then apply the estimator of Eq. (3) to the simulated data and ask how often the change is detected by the estimation procedure. The solid line gives the average estimate of ℛ_*t*_ ℛ, while the shaded lines denote 65% and and 95% of simulations (in particular, the shaded area for 65% of simulations is given by the 17.5 and 82.5 percentiles of the estimated ℛ_*t*_ across simulations, and the shaded area for 95% of simulations is given by the 2.5 and 97.5 percentiles of the estimated ℛ_*t*_’s). 10,000 Monte Carlo replications used. values of *m*_*t*_ by the recursion in Eq. (8). The estimated value of ℛ_*t*_ is then given by the estimator in Eq. (3).

The results of the power analysis are shown in Figure A.6. We observe that in 95% of the simulations, the change in ℛ_*t*_ is detected as soon as two days after the drop in ℛ_*t*_. Hence, the analysis in Section 3.3 appears sufficiently powerful to detect moderate changes in ℛ_*t*_. The key reason why the analysis has high statistical power, even though the signal-to-noise ratio is quite low (see Table A.2) is that data from multiple countries are used to obtain cross-country averages. This feature of the estimation procedure reduces estimation error substantially. While the signal-to-noise ratio is fairly low, we also note that the weight placed on data is that are more than one-week old is only (1 − *ω*)^7^ ≈ 15.2%. Hence, one week after the change in ℛ_*t*_, the estimates of ℛ_*t*_ are based primarily on data received after the change in ℛ_*t*_.

**Table A.3.**
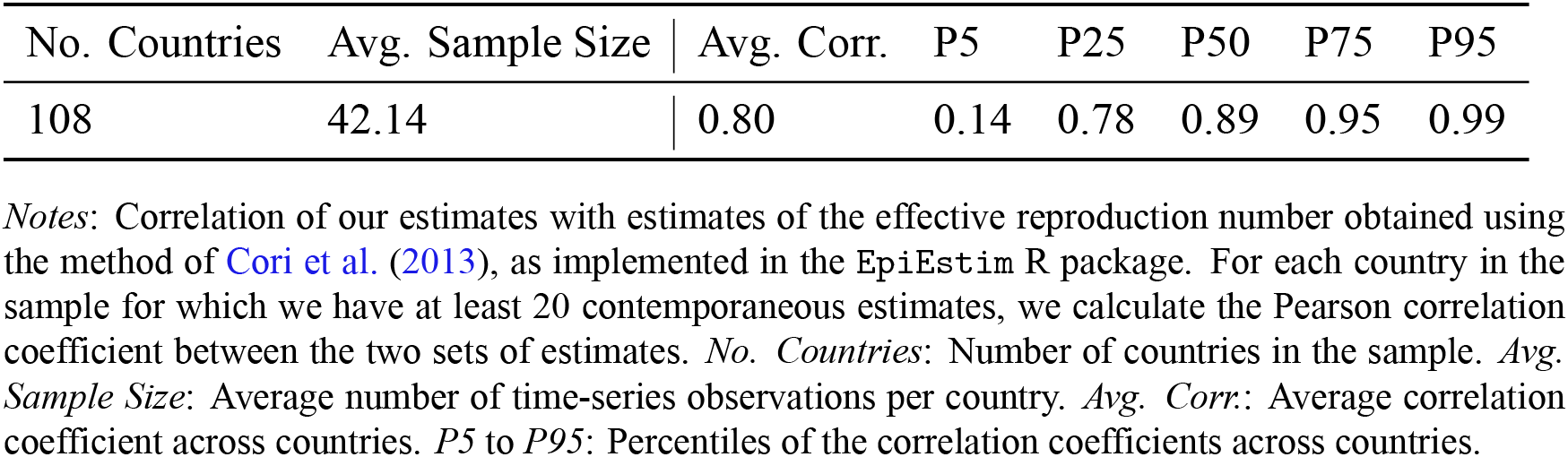
Comparison With Cori et al Estimates.

| No. Countries | Avg. Sample Size | Avg. Corr. | P5 | P25 | P50 | P75 | P95 |
| --- | --- | --- | --- | --- | --- | --- | --- |
| 108 | 42.14 | 0.80 | 0.14 | 0.78 | 0.89 | 0.95 | 0.99 |

*Notes:* Correlation of our estimates with estimates of the effective reproduction number obtained using the method of Cori et al. \(2013\), as implemented in the EpiEstim R package. For each country in the sample for which we have at least 20 contemporaneous estimates, we calculate the Pearson correlation coefficient between the two sets of estimates. *No. Countries:* Number of countries in the sample. *Avg. Sample Size:* Average number of time-series observations per country. *Avg. Corr.:* Average correlation coefficient across countries. *P5 to P95:* Percentiles of the correlation coefficients across countries.

The power analysis in the current section is arguably conservative. Specifically, we assume that after the abrupt decline, ℛ_*t*_ follows a random walk rather than staying fixed at the new level. As a result, as time goes on, the estimates of ℛ_*t*_ become more “spread out” across simulations, as is visible towards the end of Figure A.6.

### A.10 Comparison With EpiEstim Estimates

In this section, we compare our estimates of ℛ for COVID-19 with estimates obtained using the method of Cori et al. (2013); see also Thompson et al. (2019). The method proposed by Cori et al. (2013)—arguably the most widely used approach to estimating the effective reproduction number of an infectious disease—is implemented in a popular R package `EpiEstim`.

First, we download estimates of ℛ obtained using `EpiEstim` provided by Xihong Lin’s Group in the Department of Biostatistics at the Harvard Chan School of Public Health (http://metrics.covid19-analysis.org/). These estimates, just as in our case, are obtained using data from the John Hopkins CSSE repository. The parametrization used in the `EpiEstim` estimation assumes a time window of 7 days, and a gamma-distributed serial interval with a mean of 5.2 days and a standard deviation of 5.1 days. Next, we merge these estimates to our full sample of estimates. We restrict attention to countries for which we have at least 20 contemporaneous estimates from each of the two different methods. That leaves us with 108 countries and, on average, 42.14 time-series observations per country. Finally, we calculate the Pearson correlation coefficient between the two estimates of ℛ for each country.

The results of this exercise are given in Table A.3. The two sets of estimates are highly correlated, with the average correlation coefficient equal to 0.80 (median: 0.89). The interquartile range is 0.78–0.95. These findings suggest that the two estimates are highly consistent with each other, despite very different estimation methods and underlying assumptions.

## Appendix B Supplementary Figures and Tables

**Figure A.7.**
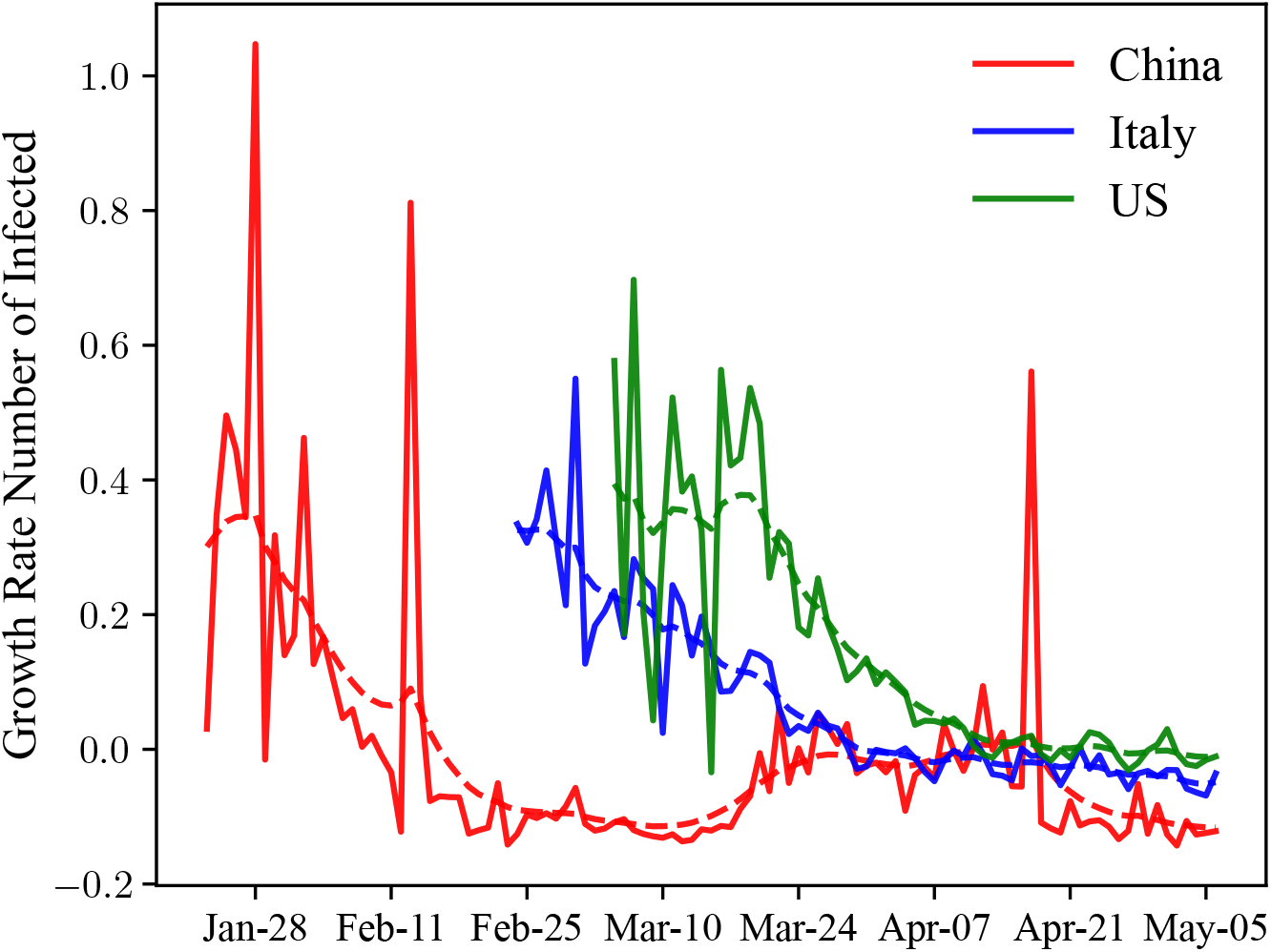
Growth Rate of the Number of Infected Individuals. *Notes*: Raw data for the growth rate of the number of infected individuals (solid lines) and our estimate of its time-varying average (dashed lines) for China, Italy, and the US.

**Figure A.8.**
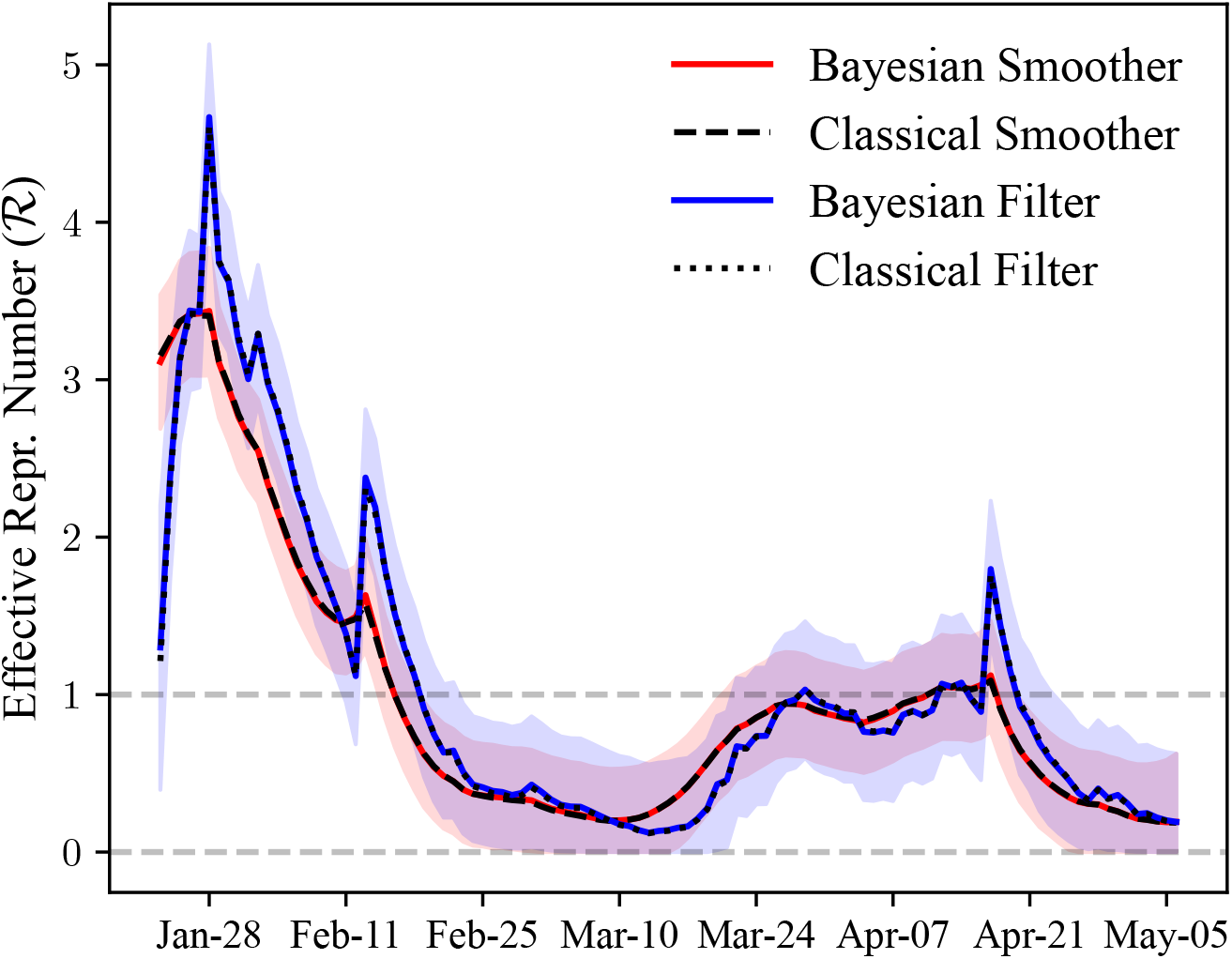
Filtered and Smoothed Estimates: China. *Notes*: Estimated effective reproduction number (ℛ) for China: filtered and smoothed estimates, using both Bayesian and classical estimation procedures. The Bayesian estimates are given by our baseline estimation procedure, as explained in the text. The classical estimates are obtained by maximum likelihood estimation (with diffuse initial conditions). The smoothed estimates use information from the full sample, while the filtered estimates at time *t* only use information up to time *t*. 65% credible bounds shown by the shaded areas.

**Figure A.9.**
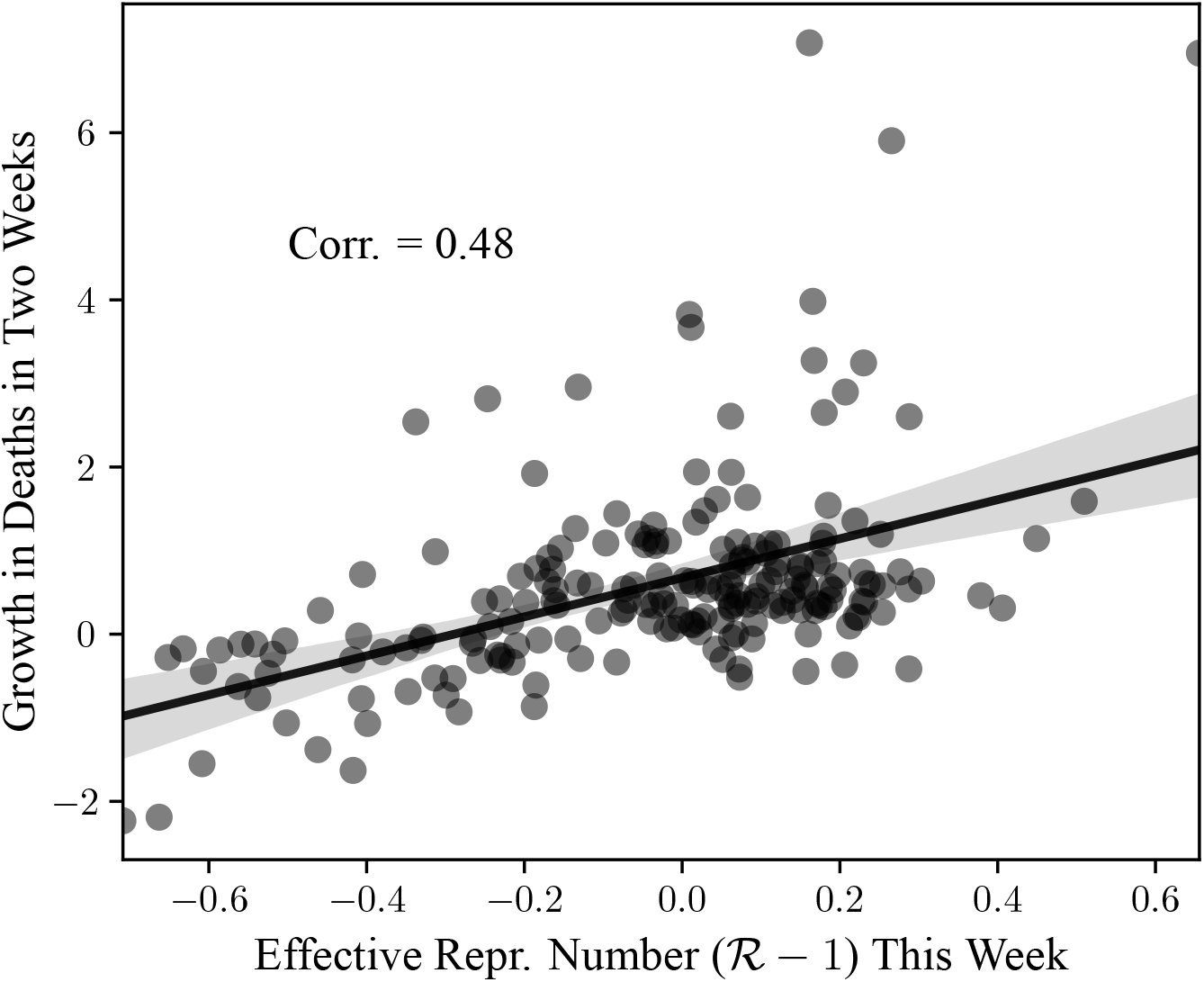
ℛ_*t*_ and Deaths in Two Weeks. *Notes*: Relationship between current estimates of the effective reproduction number (ℛ_*t*_) and the growth rate of the number of new deaths in two weeks. The data is aggregated to a weekly frequency. Both variables are residualized to subtract country fixed effects by performing the within transformation. See Figure A.4 for more details.

**Figure A.10.**
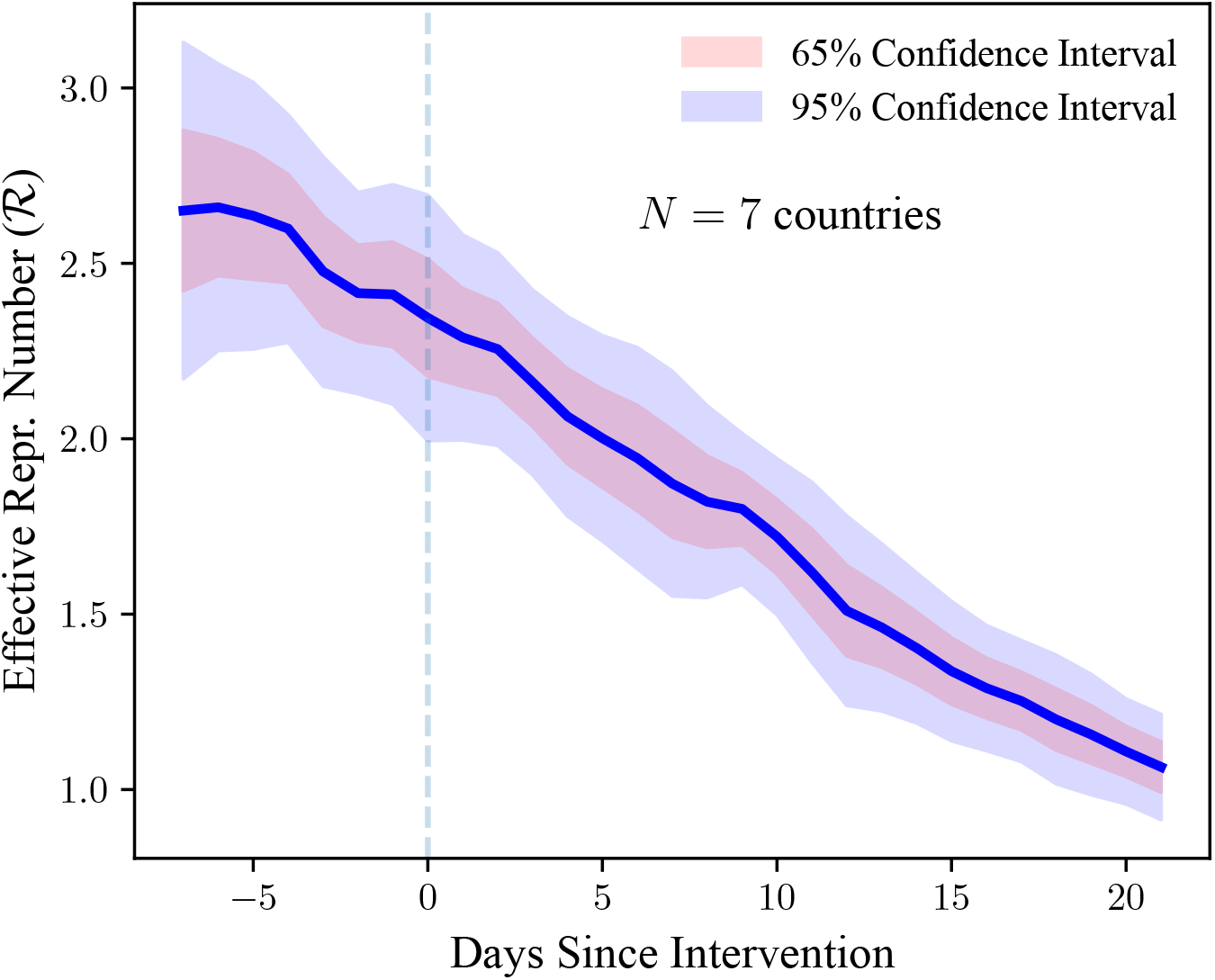
ℛ_*t*_ and Policy Interventions: Bans of Public Events. *Notes*: The graph plots the estimated effective reproduction number (ℛ_*t*_) one week before and three weeks after public events are banned in a country. The original sample consists of 14 European countries studied by Flaxman et al. (2020). For the event-study graph, we restrict the sample to countries for which data on ℛ_*t*_ is available for the whole event window. Heteroskedasticity-robust confidence bounds are shown by the shaded areas.

**Figure A.11.**
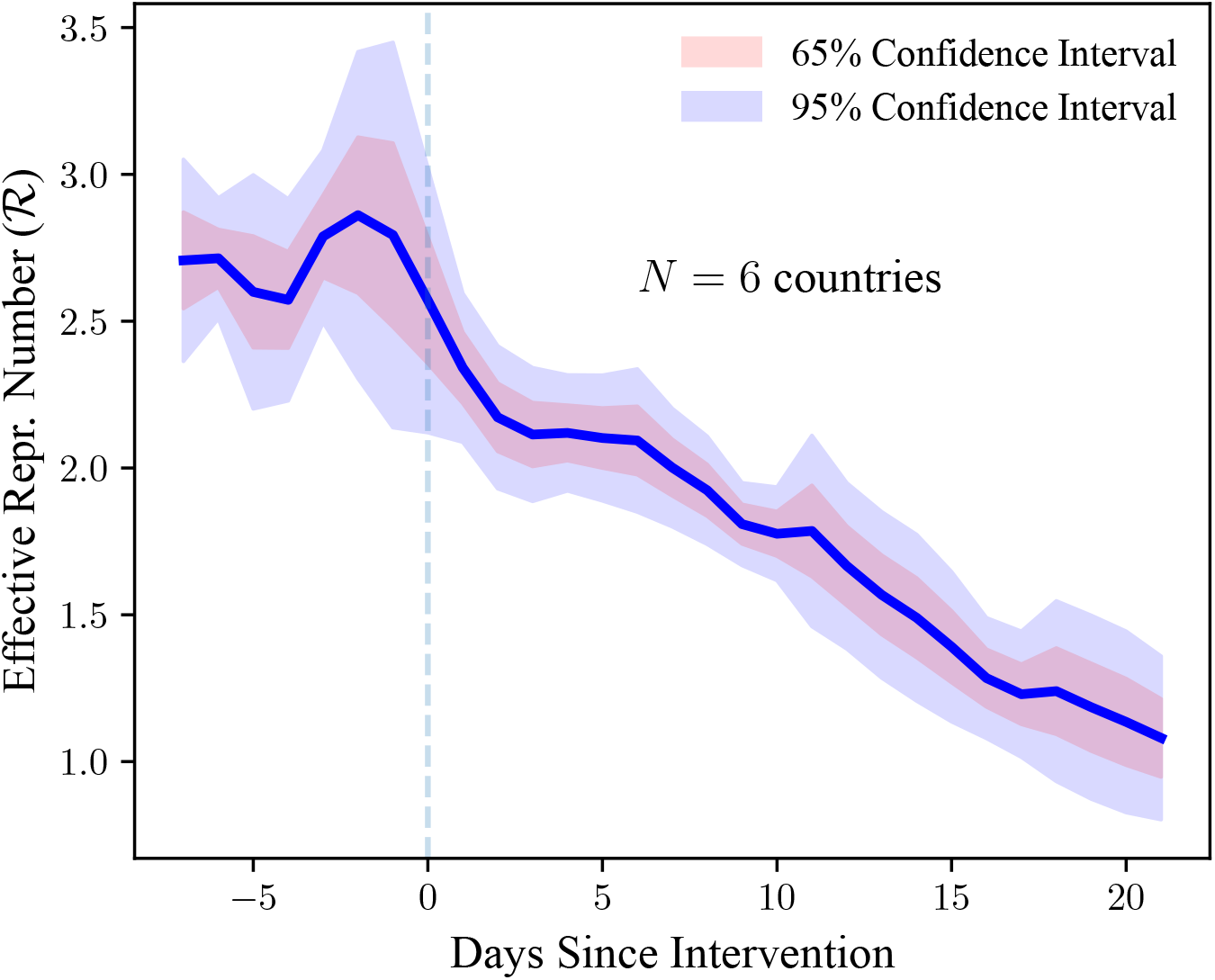
ℛ_*t*_ and Policy Interventions: Case-Based Measures. *Notes*: The graph plots the estimated effective reproduction number (ℛ_*t*_) one week before and three weeks after case-based measures are introduced in a country. The original sample consists of 14 European countries studied by Flaxman et al. (2020). For the event-study graph, we restrict the sample to countries for which data on ℛ_*t*_ is available for the whole event window. Heteroskedasticity-robust confidence bounds are shown by the shaded areas.

**Figure A.12.**
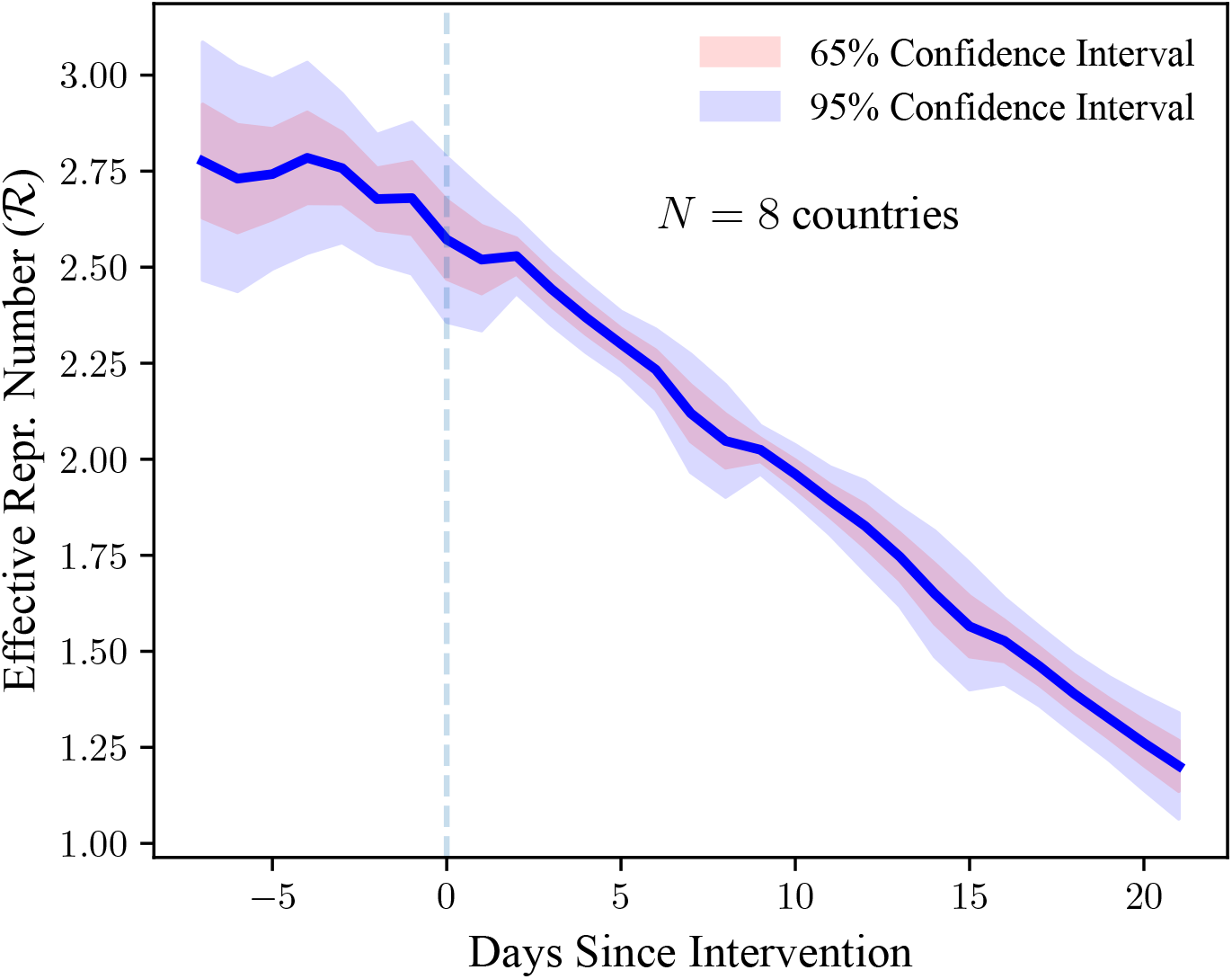
ℛ_*t*_ and Policy Interventions: School Closures. *Notes*: The graph plots the estimated effective reproduction number (ℛ_*t*_) one week before and three weeks after school closures are ordered in a country. The original sample consists of 14 European countries studied by Flaxman et al. (2020). For the event-study graph, we restrict the sample to countries for which data on ℛ_*t*_ is available for the whole event window. Heteroskedasticity-robust confidence bounds are shown by the shaded areas.

**Figure A.13.**
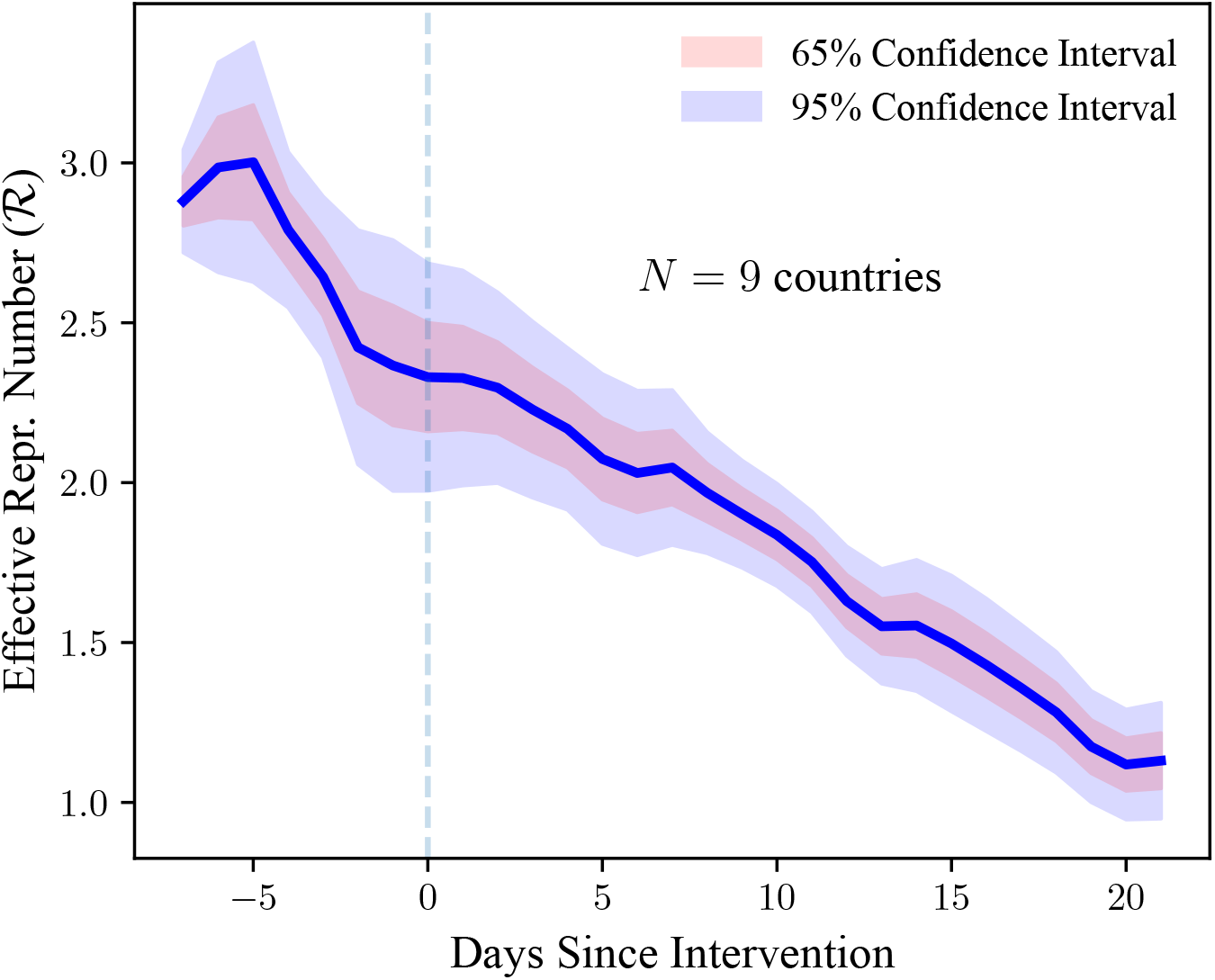
ℛ_*t*_ and Policy Interventions: Social Distancing. *Notes*: The graph plots the estimated effective reproduction number (ℛ_*t*_) one week before and three weeks after social distancing is encouraged in a country. The original sample consists of 14 European countries studied by Flaxman et al. (2020). For the event-study graph, we restrict the sample to countries for which data on ℛ_*t*_ is available for the whole event window. Heteroskedasticity-robust confidence bounds are shown by the shaded areas.

**Table A.4.**
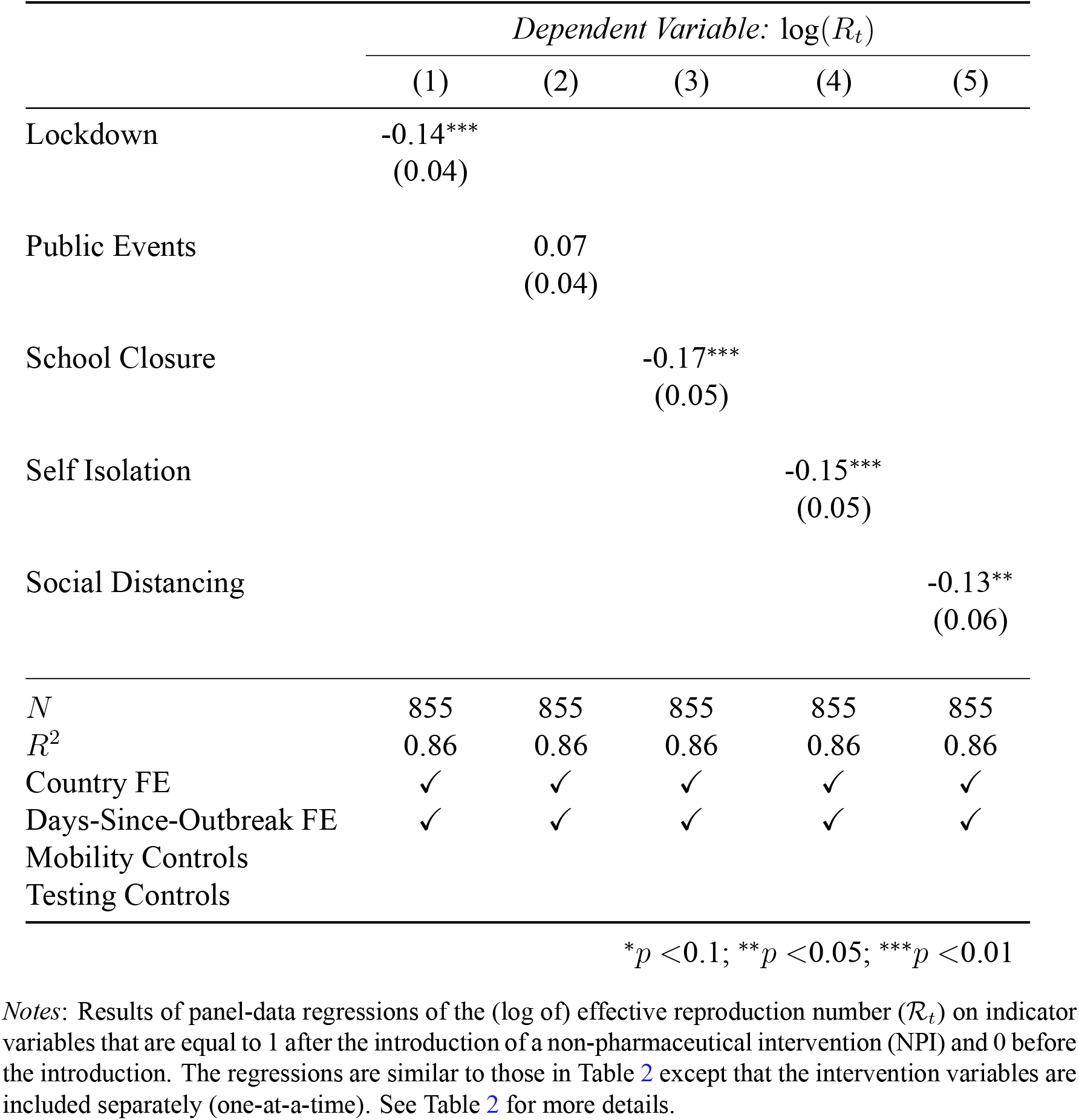
Effective Reproduction Number and NPIs.

|  | *Dependent Variable: $\log(R_t)$* |  |  |  |  |
| --- | --- | --- | --- | --- | --- |
|  | (1) | (2) | (3) | (4) | (5) |
| Lockdown | -0.14***<br>(0.04) |  |  |  |  |
| Public Events |  | 0.07<br>(0.04) |  |  |  |
| School Closure |  |  | -0.17***<br>(0.05) |  |  |
| Self Isolation |  |  |  | -0.15***<br>(0.05) |  |
| Social Distancing |  |  |  |  | -0.13**<br>(0.06) |
| *N* | 855 | 855 | 855 | 855 | 855 |
| *R*<sup>2</sup> | 0.86 | 0.86 | 0.86 | 0.86 | 0.86 |
| Country FE | ✓ | ✓ | ✓ | ✓ | ✓ |
| Days-Since-Outbreak FE | ✓ | ✓ | ✓ | ✓ | ✓ |
| Mobility Controls |  |  |  |  |  |
| Testing Controls |  |  |  |  |  |

\* $p < 0.1$ ; \*\* $p < 0.05$ ; \*\*\* $p < 0.01$
*Notes:* Results of panel-data regressions of the (log of) effective reproduction number ( $\mathcal{R}_t$ ) on indicator variables that are equal to 1 after the introduction of a non-pharmaceutical intervention (NPI) and 0 before the introduction. The regressions are similar to those in Table 2 except that the intervention variables are included separately (one-at-a-time). See Table 2 for more details.

**Table A.5.** GATHER Checklist.

| Item # | Checklist item | Reported on page # |
| --- | --- | --- |
| **Objectives and funding** |  |  |
| **1** | Define the indicator(s), populations (including age, sex, and geographic entities), and time period(s) for which estimates were made. | 6 |
| **2** | List the funding sources for the work. | TBD |
| **Data inputs** |  |  |
| *For all data inputs from multiple sources that are synthesized as part of the study:* |  |  |
| **3** | Describe how the data were identified and how the data were accessed. | 6 |
| **4** | Specify the inclusion and exclusion criteria. Identify all ad-hoc exclusions. | 7 |
| **5** | Provide information on all included data sources and their main characteristics. For each data source used, report reference information or contact name/institution, population represented, data collection method, year(s) of data collection, sex and age range, diagnostic criteria or measurement method, and sample size, as relevant. | 6 |
| **6** | Identify and describe any categories of input data that have potentially important biases (e.g., based on characteristics listed in item 5). | 26 |
| *For data inputs that contribute to the analysis but were not synthesized as part of the study:* |  |  |
| **7** | Describe and give sources for any other data inputs. | 11 |
| *For all data inputs:* |  |  |
| **8** | Provide all data inputs in a file format from which data can be efficiently extracted (e.g., a spreadsheet rather than a PDF), including all relevant meta-data listed in item 5. For any data inputs that cannot be shared because of ethical or legal reasons, such as third-party ownership, provide a contact name or the name of the institution that retains the right to the data. | 1 |
| **Data analysis** |  |  |

**Table A.5** **GATHER Checklist**
| **Item #** | **Checklist item** | **Reported on page #** |
| --- | --- | --- |
| **9** | Provide a conceptual overview of the data analysis method. A diagram may be helpful. | 4,5 |
| **10** | Provide a detailed description of all steps of the analysis, including mathematical formulae. This description should cover, as relevant, data cleaning, data pre-processing, data adjustments and weighting of data sources, and mathematical or statistical model(s). | 4,5 |
| **11** | Describe how candidate models were evaluated and how the final model(s) were selected. | 4, 5, 6, 21, 22, 23, 24, 25 |
| **12** | Provide the results of an evaluation of model performance, if done, as well as the results of any relevant sensitivity analysis. | 26, 27, 28, 29, 30, 31, 32, 33 |
| **13** | Describe methods for calculating uncertainty of the estimates. State which sources of uncertainty were, and were not, accounted for in the uncertainty analysis. | 5, 21 to 25 |
| **14** | State how analytic or statistical source code used to generate estimates can be accessed. | 1 |
| **Results and discussion** |  |  |
| **15** | Provide published estimates in a file format from which data can be efficiently extracted. | 1 |
| **16** | Report a quantitative measure of the uncertainty of the estimates (e.g. uncertainty intervals). | 8 |
| **17** | Interpret results in light of existing evidence. If updating a previous set of estimates, describe the reasons for changes in estimates. | 8,9,15 |
| **18** | Discuss limitations of the estimates. Include a discussion of any modelling assumptions or data limitations that affect interpretation of the estimates. | 15 |

*Notes:* GATHER checklist (gather-statement.org) to facilitate the evaluation and replication of our empirical analysis.

